# Decoding distinctive features of plasma extracellular vesicles in amyotrophic lateral sclerosis

**DOI:** 10.1101/2020.08.06.20169300

**Authors:** Laura Pasetto, Stefano Callegaro, Alessandro Corbelli, Fabio Fiordaliso, Deborah Ferrara, Laura Brunelli, Giovanna Sestito, Roberta Pastorelli, Elisa Bianchi, Marina Cretich, Marcella Chiari, Cristina Potrich, Cristina Moglia, Massimo Corbo, Gianni Sorarù, Christian Lunetta, Andrea Calvo, Adriano Chiò, Gabriele Mora, Maria Pennuto, Alessandro Quattrone, Francesco Rinaldi, Vito Giuseppe D’Agostino, Manuela Basso, Valentina Bonetto

**Affiliations:** Istituto di Ricerche Farmacologiche Mario Negri IRCCS, Milano (Italy); Department of Mathematics “Tullio Levi-Civita”, University of Padova; Department of Cellular, Computational and Integrative Biology – CIBIO, University of Trento, Trento, Italy; Consiglio Nazionale delle Ricerche, Istituto di Scienze e Tecnologie Chimiche “Giulio Natta” (SCITEC-CNR), Milano, Italy; Centre for Materials and Microsystems, Fondazione Bruno Kessler, Trento, Italy & Istituto di Biofisica, Consiglio Nazionale delle Ricerche, Trento, Italy; ‘Rita Levi Montalcini’ Department of Neuroscience, Università degli Studi di Torino, Torino, Italy; Department of Neurorehabilitation Sciences, Casa Cura Policlinico (CCP), Milano, Italy; Department of Neuroscience, University of Padova, 35122 Padova, Italy; NEuroMuscular Omnicentre (NEMO), Serena Onlus Foundation, Milano; Department of Neurorehabilitation, ICS Maugeri IRCCS, Milano, Italy; Department of Biomedical Sciences (DBS), University of Padova, 35131 Padova, Italy; Veneto Institute of Molecular Medicine (VIMM), 35129 Padova, Italy

**Keywords:** Extracellular vesicles, HSP90, PPIA, phosphorylated TDP-43, biomarkers, machine learning, plasma

## Abstract

**Background:** Amyotrophic lateral sclerosis (ALS) is a multifactorial, multisystem motor neuron disease for which currently there is no effective treatment. There is an urgent need to identify biomarkers to tackle the disease’s complexity and help in early diagnosis, prognosis, and therapy. Extracellular vesicles (EVs) are nanostructures released by any cell type into body fluids. Their biophysical and biochemical characteristics vary with the parent cell’s physiological and pathological state and make them an attractive source of multidimensional data for patient classification and stratification.

**Methods:** We analyzed plasma-derived EVs of ALS patients (n= 106) and controls (n=96), and SOD1^G93A^ and TDP-43^Q331K^ mouse models of ALS. We purified plasma EVs by nickel-based isolation, characterized their EV size distribution and morphology respectively by nanotracking analysis and transmission electron microscopy, and analyzed EV markers and protein cargos by Western blot and proteomics. We used machine learning techniques to predict diagnosis and prognosis.

**Results:** Our procedure resulted in high-yield isolation of intact and polydisperse plasma EVs, with minimal lipoprotein contamination. There were more particles in the plasma of ALS patients and the two mouse models of ALS while their average diameter was smaller. HSP90 was differentially represented in ALS patients and mice compared to the controls. In terms of disease progression, the levels of cyclophilin A, with the EV size distribution, distinguished fast and slow disease progressors, suggesting a new means for patient stratification. We also measured the levels of phosphorylated TDP-43 and showed that is not an intravesicular cargo of plasma-derived EVs.

**Conclusions:** Our analysis unmasked features in plasma EVs of ALS patients with potential straightforward clinical application. We conceived an innovative mathematical model based on machine learning which, by integrating EV size distribution data with protein cargoes, gave very high prediction rates for disease diagnosis and prognosis.

## Background

Amyotrophic lateral sclerosis (ALS) is a rare and fatal neurodegenerative disorder with an incidence of 3.03 cases per 100,000 persons [1]. ALS leads to selective loss of upper and lower motor neurons resulting in progressive paralysis and death within a few years from onset. About 50% of patients also develop non-motor symptoms with cognitive and behavioral changes that may appear before or after motor impairment [2]. ALS’s biggest challenge is identifying reproducible biochemical biomarkers to predict the disease in the early phase and that change during its progression. These biomarkers should reveal an adaptive response to a toxic stimulus before the degeneration starts. For example, the prodromal stage of Alzheimer’s disease is monitored well by detecting the increase of amyloid protein peptides in the brain up to ten years before the onset of symptoms [3]. In ALS, TDP-43 is a clear hallmark of the disease; it has been observed as highly post-translationally modified (cleaved and hyperphosphorylated) in protein inclusions in 97% of *post mortem* specimens from patients, but its analysis in biofluids is still hard to reproduce [4]. A promising assay to detect pathological species of TDP-43 in cerebrospinal fluid (CSF) has been recently developed [6], however methods for plasma/serum are lacking.

Neurofilaments are promising biochemical biomarkers to diagnose ALS even before the onset of clinical symptoms [7–9]. However, a drawback is that neurofilaments increase not only in ALS but in other neurodegenerative conditions too [10]. Moreover, neurofilaments are the end-product of a process of degradation in the axons. Their detection in biofluids corresponds to damage that has probably started many years before.

Extracellular vesicles (EVs) are nano-or micro-sized membranous particles released by any cells and can be found in biological fluids. There are two types of EVs, namely exosomes, that originate from late endosome/multivesicular bodies, and microvesicles that bud directly from the plasma membrane [11]. Since circulating vesicles comprise both exosomes and microvesicles, the inclusive term EVs is now preferred [12].

EVs carry specific sets of lipids, nucleic acids, and proteins, some of them common to all EVs, such as cytosolic proteins and chaperones, while others are unique and reflect specialized functions of the cell of origin [13]. In neurodegenerative diseases, there is emerging evidence that EVs might be involved in the spread of the disease since several pathogenic misfolded proteins are associated with plasma- or CSF-derived EVs, as reviewed in Basso and Bonetto, 2016 [14]. The presence of alpha synuclein, prion protein, amyloid protein precursor, and superoxide dismutase 1 (SOD1) in EVs has made them unusually attractive as a source of biomarkers.

Similarly, nearly all proteins linked to ALS have been detected in EVs from cell-conditioned media [15]. In 2013 we found that astrocytes from transgenic mice expressing SOD1 with glycine 93-to-alanine mutation (G93A) released more EVs in the culture media than controls [16]. These EVs contained SOD1^G93A^ and induced selective motor neuron death in an astrocyte-spinal neuron co-culture paradigm. TDP-43, FUS, and SOD1 have also been found in EVs isolated from ALS patients’ biological fluids [17]. Because EVs are released by damaged cells in the central nervous system (CNS) and transported to the periphery [18], they bring the CNS information to the blood. They may therefore be valuable biomarkers of ALS, especially, if their size distribution, number, and cargo could differentiate ALS patients from other conditions and stratify according to the rate of progression, e.g., fast or slow ALS. However, the purification of EVs from plasma remains a challenging task in the clinical setting and there is still no consensus on a “gold method” to isolate pure EVs in clinics [19].

We used a charge-based EV purification protocol, namely nickel-based isolation (NBI), which is low-cost and allows for a fast enrichment of vesicles [20]. With our pipeline of analysis, we unmasked certain features in plasma EVs of ALS patients that might have straightforward application in a clinical setting. Our data indicate that EVs are indeed promising biomarkers, and their parameters can be used to predict the disease and the type of progression.

## Methods

### Participants and clinical characterization

The study was approved by the ethics committees of all the centers involved in the study, Salvatore Maugeri Foundation IRCCS, Milan; NEuroMuscular Omnicentre (NEMO), Milan; Casa Cura Policlinico, Milan; and ‘Rita Levi Montalcini’ Department of Neuroscience, Università degli Studi di Torino, Turin; written informed consent was obtained from all subjects. The SBMA plasma samples were obtained by the Telethon Network of Genetic Biobank, University of Padova, Italy. The study included 106 ALS patients and 96 controls [36 healthy subjects, including plasma from 5 anonymized healthy volunteers enrolled according to the protocol 2018-008 approved by the University of Trento, 28 muscular dystrophies (MD), 32 spinal and bulbar muscle atrophy (SBMA)]. The main demographic and clinical characteristics of the subjects are listed in Table 1. ALS patients were divided into two groups according to the disease progression rate, defined by the median ΔALSFRS-R score (48 minus the ALSFRS-R score at sampling divided by the disease duration from onset to sampling): slow/intermediate-ALS with ΔALSFRS-R < 0.96 (slow-ALS) and fast-ALS with ΔALSFRS-R > 0.96. All cases were sporadic, and DNA was available from 90 of the 106 ALS patients and screened for SOD1, C9orf72, TDP-43, and FUS gene mutations. All blood samples were drawn within 18 months from the onset of the disease.

**Table 1.** Characteristics of ALS patients and controls

| Characteristics | ALS | Slow-ALS<sup>1</sup> | Fast-ALS<sup>2</sup> | HC<sup>3</sup> | MD<sup>4</sup> | SBMA<sup>5</sup> |
| --- | --- | --- | --- | --- | --- | --- |
| No. | 106 | 60 | 46 | 36 | 28 | 32 |
| Age at sampling, median (range) | 67 (42-89) | 65 (42-81) | 70 (48-89) | 61 (24-78) | 53 (25-78) | 62 (46-83) |
| Sex (male/female) | 64/42 | 41/19 | 23/23 | 15/21 | 13/15 | 32/0 |
| Site of disease onset (bulbar/spinal) | 28/77 | 7/52 | 21/25 | - | - | - |
| ALSFRS-R at sampling, median (range) | 34 (2-47) | 37 (4-35) | 27 (2-41) | - | - | - |
| ΔALSFRS-R<sup>6</sup>, median (range) | 0.86 (0.05-9.0) | 0.54 (0.05-0.95) | 1.48 (0.97-9) | - | - | - |
| Disease duration<sup>7</sup>, median months (range) | 14 (3-179) | 15 (3-179) | 13 (3-46) | - | - | - |
| Gene mutations (SOD1/C9orf72/TDP-43/FUS) | 10/106 (4/5/1/0) | 6/60 (3/2/1/0) | 4/46 (1/3/0/0) | - | - | - |

<sup>1</sup>Slow-ALS: ΔALSFRS-R < 0.96; <sup>2</sup>Fast-ALS: ΔALSFRS-R > 0.96; <sup>3</sup>HC: Healthy control; <sup>4</sup>MD: muscular dystrophy; <sup>5</sup>SBMA: spinal and bulbar muscular atrophy; <sup>6</sup> ΔALSFRS-R: 48– ALSFRS-R score at the plasma collection/time between symptom onset and sampling; <sup>7</sup>Disease duration: from symptom onset to plasma collection.

### Animal model

Mice were maintained at 21±1 °C with relative humidity 55%±10% and 12 hours of light. Food (standard pellets) and water were supplied *ad libitum*. All the procedures involving animals and their care carried out at the Mario Negri Institute for SOD1^G93A^ mice and at the CIBIO University of Trento for TDP-43^Q331K^ mice were conducted as described by the institutional guidelines, that are in accordance with national (D.L. no. 116, G.U. suppl. 40, February 18, 1992, no. 8, G.U., 14 July 1994) and international laws and policies (EEC Council Directive 86/609, OJ L 358, December 12,1987; National Institutes of Health Guide for the Care and Use of Laboratory Animals, US National Research Council, 1996). The mice were bred in conventional specific pathogen-free mouse facilities.

The SOD1^G93A^ mouse line on a homogeneous 129S2/SvHsd background derives from the B6SJL-TgNSOD-1-SOD1G93A-1Gur line originally obtained from The Jackson Laboratory (Bar Harbor, ME, USA); it expresses about 20 copies of mutant human SOD1^G93A^ [21]. The use of SOD1^G93A^ mice was authorized in protocol no. 657/2018-PR.

TDP-43^Q331K^ transgenic line 103 mice (stock 17033) express a myc-tagged, human TAR DNA binding protein carrying the ALS-linked Q331K mutation (huTDP-43^Q331K^) directed to brain and spinal cord by the mouse prion protein promoter on the C57BL/6NJ background. The expression of the protein is 1.5 times that of the endogenous TDP-43 [22]. The use of TDP-43^Q331K^ mice was authorized in protocol no. 603/2017-PR.

Mice were deeply anesthetized with ketamine hydrochloride (IMALGENE, 100 mg/kg; AlcyonItalia) and medetomidine hydrochloride (DOMITOR, 1 mg/kg; Alcyon Italia) by intraperitoneal injection and blood was drawn and centrifuged to isolate plasma, as described in ‘Blood sampling’. SOD1^G93A^ female mice were analyzed at 10 and 16 weeks of age (SOD1^G93A^ 10 weeks=4; SOD1^G93A^ 16 weeks=5), corresponding to the pre-symptomatic and symptomatic stage of disease. Male TDP-43^Q331K^ mice were analyzed at 10 months, corresponding to the symptomatic stage of the disease (TDP-43^Q331K^ 10 months=7)[22].

The corresponding age-matched nontransgenic mice were used as controls for SOD1^G93A^ (n=10) and TDP-43^Q331K^ mice (n=8). Genotyping for SOD1^G93A^ and TDP-43^Q331K^ was done by standard PCR using primer sets designed by The Jackson Laboratory. The number of animals was calculated on the basis of experiments designed to reach a power of 0.8, with a minimum difference of 20% (α=0.05).

### Blood sampling and plasma isolation

Samples of peripheral venous blood from patients and controls were collected in EDTA pre-coated vials (BD Vacutainer K2EDTA). Blood was centrifuged at 3000 x g for 20 minutes, frozen and kept at -80 °C until further analysis.

For animal models, up to 500 µL of blood per mouse was sampled by intracardiac puncture, collected in EDTA pre-coated vials and centrifuged at 3000 x g for 10 minutes. Mouse plasma samples were stored at -80 °C until EV isolation. Only samples frozen once were used in the analyses.

### EV isolation

#### Ultracentrifugation (UC)

Plasma samples (500µL) were diluted with an equal volume of PBS and subjected to UC to remove cells, dead cells and cellular debris (10 min at 200 x g, 10 min at 1000 x g and 25 min at 20,000 x g at 4 °C) (rotor type JA 25.50), as already reported [16]. The final supernatant was then ultracentrifuged for 1 hour at 100.000 x g (rotor type 70i) at 4 °C to pellet EVs. The pellet was washed twice by suspension in PBS and ultracentrifugation for 1 hour at 100,000 x g at 4 °C to eliminate contaminating proteins. EVs were analyzed within one week from extraction.

#### Nickel-based isolation (NBI)

Human (500µL) and mouse plasma (100µL) were centrifuged to remove cells, dead cells and cellular debris (10 min at 2800 x g at RT) and the supernatant was diluted in filtered PBS respectively one and ten times. The diluted plasma was incubated with 25 µL/mL of nickel-functionalized agarose beads [23] and placed in orbital shaking for 30 min at RT. The beads with the bound EVs were gently centrifuged (5 min at 1000 x g at RT) and the supernatant was discarded. Elution buffer (PBS, EDTA 3.2 mM, NaCl 2 mM, citric acid 45 µM) [23] (100 µL/mL) was added to the beads and incubated in a Thermomixer (10 min at 750 rpm at 28 °C). The beads were pelleted (1 min at 600-800 x g at RT) and the supernatant-containing EVs-was transferred to a new tube. The starting plasma volume per subject was not more than 500 µl and was enough for the analysis of the physical and biochemical EV parameters described. EVs were analyzed within one week from extraction.

### Transmission electron microscopy (TEM)

Five µL drops of isolated EVs in PBS were left to dry at room temperature for 30 min on a 100 mesh formvar/carbon coated copper grid (EMS, Hatfield, PA, USA) and fixed with 4% paraformaldehyde and 2 % glutaraldehyde in 0.12 M phosphate buffer (pH 7.4) for 30 min. EVs were then postfixed in OsO_4_ 1% in 0,12 M cacodylate buffer (pH 7.4) for 30 min and counterstained with uranyl acetate (oversatured solution) for 15 min. After dehydration by a graduated scale of ethanols the grids were embedded in LR White. For immuno-electron microscopy, five µL drop of EVs were placed to dry at room temperature for 30 min on formvar/carbon coated nikel grid (EMS, Hatfield, PA, USA) and fixed with 4% paraformaldehyde and 0.25 % glutaraldehyde in 0.12 M phosphate buffer (pH 7.4) for 30 minutes. EVs were then incubated with a mouse anti-phospho TDP-43 (pS409/410) (1:100 dilution, Cosmo Bio Co., Ltd.) overnight at 4°C, followed by a goat anti-mouse antibody conjugated to a 12 nm colloidal gold (1:70 dilution, Jackson Immunoresearch) in block solution (BSA 0.5%) for 45 minutes at 37 °C. After post-fixation with 2% glutaraldehyde, samples were counterstained with uranyl acetate and embedded in LR White. Grids were observed with an Energy Filter Transmission Electron Microscope (EFTEM, Zeiss Libra® 120) equipped with an yttrium aluminium garnet (YAG) scintillator slow-scan CCD camera (Sharp eye, TRS).

### Nanotracking analysis (NTA)

NTA was carried out to detect the size distribution and concentration of isolated EVs using a NanoSight NS300 (equipped with a sCMOS camera and 532 nm diode laser; Malvern scientific). Data were acquired and processed by two operators throughout the study, using NTA software version 3.00, on the basis of manufacturer’s recommendations. Before starting the analysis, samples were centrifuged at 10,000 x g for 5 min to remove remaining beads. Human and mouse samples were then diluted in filtered PBS respectively 100 and 50 times (final volume of 1mL), to maintain 20-40 particles per field of view and 5 × 60 s videos were recorded (at camera level 11-12); for analysis, at least 1,000 completed tracks were required per measurement. Analyses were always carried out at the same settings (detection limit 3). To assess the quality of the analysis, for each sample we considered the ratio between the total particle tracks and the valid particle tracks. The analysis was considered valid when the ratio was less than 5. The quantification of EVs is described throughout per mL. NTA analysis was also used to provide the mean and mode with 95% CI. The D10, D50 and D90, which are the size points below which 10%-50% and 90% of the particles are contained, were also considered.

We stratified EV populations as small or big EVs on the basis of a cut-off of 130 nm, which is the median D50 of all the samples analyzed. For each sample a distribution curve with the row data of NTA was generated and set with the same x (from -2.0E+06 to 1.6E+07 no. of particles/mL) and y axis (from 0 to 400 nm) axis, and the same sizes (39.38 cm x 30.43 cm). The relative abundance of small versus big EVs was calculated from the area under the curve (AUC) for size distribution curve, below or above the 130 nm cut-off. The AUC was measured with ImageJ software [24].

Sample extraction was randomized for human and mouse plasma. The operators were blinded during the NTA.

### Tunable resistive pulse sensing (TRPS)

TRPS measurements were made with a gold qNano instrument (Izon Ltd.) mounting a polyurethane nanopore membrane NP200 (analysis range 85-500 nm) and NP400 (analysis range 185-1100 nm) (Izon Ltd). The electrolyte solution consisted of filtered PBS with Primo Syringe Filters 30 mm-PES membrane 0.22 µm.

### Zeta potential

Z–potential measurements were made at 25 °C with a Zeta Sizer instrument (Nano-ZS, Malvern Instruments). Samples were introduced into DTS1070 capillary cells (Malvern Panalytical) with the diffusion barrier technique, while data were analyzed with Zetasizer Software. Phase Analysis Light Scattering (PALS) was used to determine the average zeta potential of vesicles dispersed in PBS. A fast measurement process (fast field reversal mode: FFR) was selected, because of the high ionic strength of the dispersing media (PBS). Sizes were also measured before and after the zeta potential measurements to check the sample had not changed because of the measurement.

### EV-like liposomes

EV-like liposomes were prepared from a mixture composed of 20% mol egg phosphatidylcholine, 10% mol egg phosphatidylethanolamine, 15% mol dioleoylphosphatidylserine, 15% mol egg sphingomyelin, 40% mol cholesterol (adapted from [25,26]). All phospholipids were acquired from Avanti Polar Lipids. Lipid films were formed by removing the organic solvent (chloroform) from a lipid solution by rotary evaporation and vacuum drying for at least 30 min. Lipids at a final concentration of 1 mg/mL were swollen in PBS and vortexed vigorously to give multilamellar liposomes, which were then exposed to six cycles of freezing and thawing. EV-like liposomes were obtained by extruding the suspension of multilamellar liposomes with a two-syringe extruder (LiposoFast Basic Unit, Avestin Inc.). Thirty-one passages were done through two stacked polycarbonate filters (Millipore) with pores of 50 or 100 nm nominal average diameters. Finally, size and Z-potential of EV-like liposomes were measured with a Zeta Sizer instrument (Nano-ZS, Malvern Instruments).

### Proteomics

Three aliquots (0.5 mL) of a pool of plasma samples from six healthy subjects were isolated independently by UC (samples #1UC, #2UC, #3UC) and NBI (samples #1NBI, #2NBI, #3NBI). EV proteins were extracted using RIPA buffer (150 mM NaCl, 1.0% Triton, 0.5% sodium deoxycholate, 0.1% SDS, 50 mM Tris, pH 8.0). An equal amount of proteins for each sample (9 μg) was separated by 1D 4-12% Nupage Novex Bis Tris Gel (Invitrogen), stained with Bio-Safe Coomassie (Bio-Rad Laboratories) and digested with trypsin, using a published procedure [27]. Two µL of each sample were analysed on a Biobasic 18 column (150 × 0.18 mm ID, particle size 5 µm, Thermo Scientific) coupled with Q-Exactive (Thermo Scientific) via a DESI Omni Spray (Prosolia) used in nanospray mode. Peptides were eluted with a 240 min gradient of 5%-60% buffer B (80% acetonitrile) at a flow rate of 2 μL/min. The Q-Exactive was operated in data-dependent mode with a survey scan range of 400-2000 m/z and resolution 70’000 in parallel with low-resolution MS/MS scans of the 20 most abundant precursor ions with a charge ≥ 2. Dynamic exclusion of sequenced peptides was set to 15 s to reduce the number of repeated sequences. Data were acquired using Xcalibur software (Thermo Scientific). MaxQuant software (version 1.6.2.3) was used to analyze MS raw files [28]. MS/MS spectra were searched against the human Uniprot FASTA database (Version 2016) and a common contaminants database (247 entries), using the Andromeda search engine [28]. Cysteine carbamidomethylation was applied as fixed and methionine oxidation as variable modification. Enzyme specificity was set at trypsin with a maximum of two missed cleavages and a minimum peptide length of 7 amino acids.

A false discovery rate (FDR) of 1% was required for peptides and proteins. Peptides were identified with an allowed initial precursor mass deviation of up to 7 ppm and an allowed fragment mass deviation of 20 ppm. Protein identification required at least one unique peptide. A minimum ratio count of 1 was required for valid quantification events, with MaxQuant’s Label Free Quantification algorithm (MaxLFQ). Data were filtered for common contaminants and peptides only identified by side modification were excluded from further analysis.

Bioinformatic analysis was done in the Perseus software environment [29]. Protein abundance changes were computed on LFQ peak intensities. Statistical analysis was done with the non-parametric Wilcoxon-Mann-Whitney test, using p < 0.05 as cut-off (JMP Pro13 statistical software). Functional enrichment analysis was done with STRING (https://string-db.org/), using the Gene ID of the identified proteins.

### Protein extraction for EVs

Proteins from EVs isolated by NBI were precipitated with three volumes of acetone for 2 hours at 4°C with agitation, and centrifuged at 9,000 x g for 5 min at 4 °C. Pellets were suspended in 1% boiling SDS and analyzed by Western blot analyses. The pellets of EVs isolated by UC were also suspended in 1% boiling SDS and analyzed. Proteins were quantified by the BCA protein assay (Pierce).

### Antibodies

Antibodies for immunoblot (Western) were as follows: mouse monoclonal anti-cytochrome C (1:500 dilution; BD Biosciences; RRID: AB_396417); mouse monoclonal anti-calnexin (1:1000 dilution; Abcam; RRID: AB_11178981); rabbit monoclonal anti-syntenin (1:2500 dilution; Abcam; RRID: AB_11160262); rabbit monoclonal anti-GM130 (1:2500 dilution; Abcam; RRID: AB_880266); rabbit monoclonal anti-CD81 (1:1000 dilution; Cell Signaling; RRID: AB_2714207); mouse monoclonal anti-flotillin-1 (1:500 dilution; BD Transduction Laboratories; RRID: AB_398139); rabbit polyclonal anti-cyclophilin A/peptidyl-prolyl cis-trans isomerase A (PPIA) (1:5000 dilution; Proteintech; RRID: AB_2237516); rabbit polyclonal anti-HSP90 (1:1000 dilution; Enzo Life Sciences; RRID: AB_2039287); rabbit polyclonal anti-TDP-43 (1:2500 dilution; Proteintech; RRID: AB_2200505); rabbit polyclonal anti-TDP-43 (1:2500 dilution; Proteintech; RRID: AB_615042); mouse monoclonal anti-human phospho Ser409/410 TDP-43 antibody (pTDP-43) (1:2000 dilution; Cosmo Bio Co., LTD; RRID: AB_1961900); goat polyclonal anti-apoliprotein B (1:500; Abcam, RRID:AB_305987); goat polyclonal anti-apoliprotein AI (1:500 dilution; Abcam, RRID:AB_2289632); goat anti-mouse or anti-rabbit peroxidase-conjugated secondary antibodies (respectively 1:20000 and 1:10000 dilution, GE Healthcare).

### Western blot (WB)

For WB, human and mouse samples (40 μg) were separated in 12% SDS-polyacrylamide gels and transferred to polyvinylidene difluoride membranes (Millipore). For protein characterization of human EVs, to minimize inter-assay variability each blot was loaded to hold one healthy control, three ALS patients, one MD and one SBMA control, with a total of 36 blots for each protein. For mouse EVs protein analysis, to minimize inter-assay variability, each blot was loaded to hold a SOD1^G93A^ sample at 10 and 16 weeks of age with the respective controls, and a TDP-43^Q331K^ sample with relative control, with a total of six blots for each protein. In addition, for both human and mouse EVs the respective internal standards (IS) were used for all the blots to favor inter-assay analysis. The IS is a pool of human or mouse plasma EV samples. To minimize intra-assay variability, both human and mouse aliquots were prepared the same day for all the proteins analyzed and samples were loaded alternately in the blot. WB membranes were blocked with 3% (w/v) BSA (Sigma-Aldrich) and 0.1% (v/v) Tween 20 in Tris-buffered saline, pH 7.5, and incubated with primary antibodies, then with peroxidase-conjugated secondary antibodies. Blots were developed with Luminata™ Forte Western Chemiluminescent HRP Substrate (Millipore) on the ChemiDoc™ Imaging System (Bio-Rad). Densitometry was done with Image Lab software 6.0. Immunoreactivity was normalized to the Red Ponceau staining (Fluka) and to the immunosignal of the IS of each membrane. Data are expressed as arbitrary units (A.U.). Censored data were replaced with L/√2, where L is the lowest value detected in all samples.

### ELISA pNf-H

pNf-H was measured in human plasma with an ELISA kit for the human protein (EUROIMMUN #EQ-6561-9601). Censored data were replaced with L/√2, where L is the limit of detection of the assay.

#### Machine learning

In view of the limited number of samples and the presence of unbalanced classes, we built and compared two different machines learning frameworks, that is the basic one, represented in Fig. 6a, and the advanced one, in Fig. 6b. Both frameworks are divided into three different steps: data handling (see detailed description in section “Data preprocessing and oversampling”), training of the models (see detailed description in sections “Classification models” and “K-fold cross validation”) and testing of the models on an independent cohort (see details in section “Classification accuracy of prediction models”). The main difference between the two frameworks is in the first part, that is data handling. Indeed, in the basic one we just perform some simple preprocessing to clean up the training data, while in Fig. 6b, we further use some tailored oversampling strategy that enables us to better handle unbalanced training data, thus getting better performances in the end. Model training, internal validation and testing were done on the NTA curve distributions and on the data on HSP90 and PPIA values for all the EV plasma. We used Python 3.7.4 downloaded at https://www.python.org/downloads/release/python-374/ running in a Windows 10 PC x64, with Intel(R) Core(TM) i5-4200M CPU with 2.5 GHz and 4 GB of RAM. To write this Python code we used PyCharm version 2019.2, an IDE (Integrated Development Environment) developed by JetBrains https://www.jetbrains.com/pycharm/download/other.html. We embedded some Scikit-learn tools into our software. Scikit-learn (https://scikit-learn.org/stable/) is a Python open-source library that embeds simple, efficient tools for predictive data analysis and machine learning. We also developed a specific software library for analysis of the NTA curve distributions (https://github.com/tety94/rbfn, tag v1.0.0).

#### Distribution Curve Compression

As a preliminary step we normalized all the distribution curves (Supplementary Fig. 3f). Then we used machine learning tools to compress the signal. Specifically, we considered a distribution curve as a set of noisy samples picked from an unknown function, and approximated it with a fixed number of Radial Basis Functions (RBFs) [30]. We used 30 RBFs in the experiments. An example of an approximated curve is reported in Supplementary Fig. 3g. The reconstructed curve fits the original one very well. Since the coefficients related to the RBF model represent an implicit description of the approximated signal, we used them as the set of features that map the curve into a smaller dimensional space [31].

#### Data Preprocessing and oversampling

As a first step we had to preprocess and suitably split the samples in two independent sets, that is the training and the test set (respectively indicated with TR and TS in both Fig. 6a and 6b). When dealing with the dataset that includes the biomarkers, we had some missing values so, in order to impute those missing data, we used feature average when needed. In view of the limited number of samples and the presence of unbalanced classes, we built and compared two different learning machines. As mentioned above, the first one was simply obtained by using the original training data (indicated with TR in Fig. 6a). The second one was instead built up using oversampled data (indicated with OTR in Fig. 6b). We used a synthetic oversampling algorithm called MWMOTE [32] and a specific procedure to avoid overoptimistic predictions (see Fig. 6b), similar to the one proposed in [33]. This approach allowed us to avoiding overfitting, which might occur when oversampling is used the wrong way.

#### Classification models

The distribution curve features were used to feed a trained learning machine to distinguish ALS patients from others. Since our dataset consists of input-output samples, we used a supervised training approach. Supervised learning techniques are specific algorithms that train a predictor using data that consist of inputs paired with the correct outputs. We used Random Forests to build up our classification models. We used binary classification models to compare ALS with the other diseases (MD and SBMA) and HC. We also split ALS patients into two groups according to the disease progression rate (fast vs slow ALS) and built up classification models using those data. In addition to the distribution curve features, we included specific biomarkers to check for any improvement in the performance of the classifiers. We built models to compare ALS with one of the other classes based on plasma EV size distribution and HSP90 amount. We used EV size distribution and PPIA amount to distinguish fast from slow ALS.

#### K-fold cross validation

Cross validation is a resampling procedure used to evaluate machine learning models on limited data samples. The k refers to the number of groups that a given data sample is to be split into. In our tests, k = 5. A k-fold cross validation strategy was used to assess the performances of the different learning machines [34] and select the best parameter settings for the models. We chose a stratified algorithm as the generated folds preserve the percentage of samples for each class.

#### Classification accuracy of prediction models

In order to assess the performances of our prediction models, we used an independent cohort as a test set in both machine learning frameworks (see TS in Fig 6a and 6b) and considered two different measures: Precision (the fraction of relevant instances among the retrieved instances) and Recall (the fraction of relevant instances that were actually retrieved). Precision-recall curves were plotted to display performances of the different classifiers. Those curves show the trade-off between precision and recall for different thresholds. Our target/positive class in the experiments is the ALS patients.

### Statistical analyses

Different EV parameters were compared between ALS patients, MD, SBMA and HC, using one-way ANOVA. For parameters showing a significant (p < 0.05) difference in the distribution among the four categories, pairwise comparisons were made using the Dunnett’s multiple comparisons *post hoc* test, to identify the categories showing the difference. Pairwise comparisons were limited only to comparisons involving ALS patients (ALS versus MD, ALS versus SBMA, ALS versus HC). Significant and differentially expressed EV parameters between fast and slow progressing ALS patients were selected using Student’s *t* test, with p < 0.05 as cut-off. The same analyses were used to compare the EV parameters in the two isolation procedures NBI and UC. Different EV parameters were compared between controls and SOD1^G93A^ mice at 10 and 16 weeks of age, using one-way ANOVA. For parameters showing a significant (p < 0.05) difference in the distribution among the three categories, pairwise comparisons were made using the Tukey’s multiple comparisons post hoc test. Significant and differentially expressed EV parameters between TDP-43^Q331K^ and controls mice were selected using Student’s *t* test, with p < 0.05 as cut-off. Univariable and multivariable linear models were used to assess if EV parameters were associated with demographic (age and sex) and clinical characteristics of ALS patients (ALSFRS-R, site of onset, disease duration, LALSFRS-R).

## Results

### High-yield isolation of intact, pure and polydisperse plasma EVs by NBI

To test whether ALS plasma EVs could be employed as biomarkers of disease diagnosis and progression, we looked for EV isolation methods that would be fast and reproducible. We isolated plasma EVs by NBI, a cost-effective procedure that exploits the net charge of membrane vesicles in a physiological pH [20] (Fig. 1a) to capture them in a short turn-round time (<1 hour) [35]. Recovered EVs were characterized according to the guidelines of the International Society of Extracellular Vesicles [12,36]. EV samples were analyzed by immunoblotting for the presence of three positive markers (flotillin-1, syntenin, and CD81) and the absence of three negative markers (GM130, calnexin, and cytochrome C). All positive markers were enriched in EV samples compared to EV-depleted plasma, and there were no negative markers (Fig. 1b). Isolation of EVs was confirmed by TEM, which detected mostly rounded membrane particles in the 23-150 nm size range (Fig. 1c), and by TRPS, which analyzed the concentration and diameter of the particles (Supplementary Fig. 1c-f). The sizes of EVs varied, confirming that NBI recovers polydisperse particles [20].

**Figure 1.**
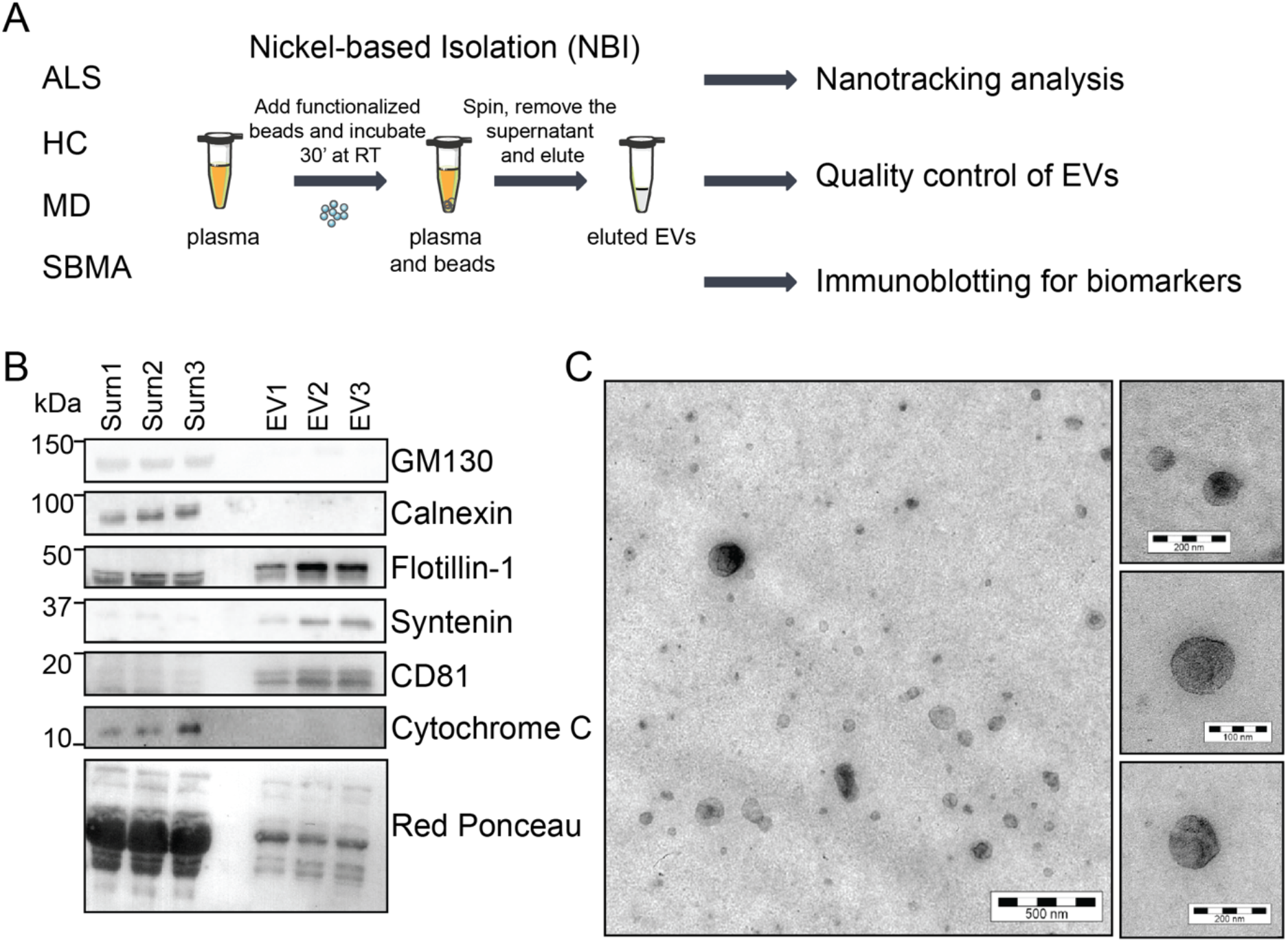
Plasma EVs are efficiently purified by the nickel-based isolation method (NBI). **a** Schematics of the purification strategy and validation. **b** Immunoblot for flotillin-1, syntenin, CD81 (markers of EVs) and GM130, calnexin and cytochrome C (contaminants of EVs) in plasma EV samples (EV1, EV2, EV3) isolated by NBI, and EV-depleted plasma (Surn1, Surn2, Surn3). Loading control represented by Red Ponceau. **c** Transmission electron microscopy (TEM) analysis of the plasma EV purified with NBI: one representative image from three independent experiments. Bar, 500 nm in the main panel; 200 nm, 100 nm, 200 nm in the insets from top to bottom.

We assessed the performance of NBI in comparison with UC, which is one of the most widely used methods with relatively high yield and purity, to isolate EVs [35,37]. We used equal volumes of plasma from 15 subjects with ALS, processed by both methods in parallel following well established protocols [13,23,38]. We analyzed the EV markers and performed the TEM for plasma EVs purified with UC (Supplementary Fig. 1a-b). Then, we measured the amount of particles/mL with the TRPS; NBI recovered respectively four and 1.5 times more particles than UC when small and big vesicles were measured, (NP200 measured show 5.2E+09 ± 6.9E+08 versus 1.2E+09 ± 1.6E+08 particles/mL; NP400 detected 8.8E+07 ± 1.2E+07 versus 5.7E+07 ± 1.1E+07) (Supplementary Fig. 1c-d).

We calculated the particles-to-µg of proteins ratio as a measure of protein contamination, and NBI gave a significantly higher value than UC (123.2 ± 9.9 versus 51.8 ± 6.5), indicative of a lower level of protein contamination (Supplementary Fig. 1g) [39]. In our settings, EVs isolated by NBI had on average a smaller diameter than those isolated by UC (NP200 recorded 84.4 nm ± 1.7 versus 98.5 nm ± 1.0; NP400 gave 210.5 nm ± 3.8 versus 275.5 nm± 7.4) (Supplementary Fig. 1e-f) likely due to a difference in the pre-clearing protocols that favors the selection of different EV populations [40]. To evaluate the NBI reproducibility, we generated liposomes with an EV-like lipidic composition (EV-like liposomes), and we diluted 1.0E+10 particles/mL in one mL of EV-depleted plasma. We ran three independent experiments in which we purified EV-like liposomes by either NBI or UC. We quantified the amount and size of each replicate by TRPS. As shown in Supplementary Fig. 1h, the EV-like liposomes purified with NBI had similar size-distribution curves among replicates (CV 8.9%). The average mean and mode diameters were 140 ± 11 and 102 ± 4 for the bigger and smaller EVs, respectively. We confirmed a similar level of reproducibility in the plasma EV samples (Supplementary Fig. 1i).

We did label-free proteomic profiling to assess whether NBI was enriching for EVs and compared NBI and UC isolated EVs. We identified 107 proteins, 60 of them overlapping (∼56% of the total) between NBI and UC samples (Supplementary Table 1 and Supplementary Fig. 2a). With both purification strategies about 70% of identified proteins were annotated as extracellular vesicle components (GO cellular component, Supplemental Table 2). Twenty-four proteins were differentially enriched either in NBI or UC (p < 0.05, Wilcoxon-Mann-Whitney test). Interestingly, 7 out of the 24 were apolipoproteins, which were enriched in UC. Lipoprotein particles are very abundant in blood and biophysically very similar to EVs; therefore they co-isolate in standard isolation procedures, and are considered contaminants in the EV analysis [41–43]. We detected twelve apolipoproteins in the UC-EVs, but only Apo-AI, Apo-AII, and Apo B-100 in the NBI-EVs, and there were significantly fewer than UC-EVs (Table 2). We also validated the decreased amount of co-purified Apolipoprotein B and AI in NBI compared to UC purification by immunoblotting (Supplementary Fig. 2c and d). Overall, the NBI method gave an 86% reduction in apolipoprotein content, indicating that it minimizes the lipoprotein particle co-isolation issue. Heat shock cognate 71 kDa protein (HSC70) and Heat shock protein HSP 90-alpha (HSP90A), two EV luminal markers, were instead, significantly higher in NBI-EVs, confirming that NBI enriches rather pure EVs (Supplementary Fig. 2b and Supplementary Table 1).

**Table 2.** Differential expression data for apolipoproteins in EV samples isolated by UC or NBI

| **Protein name** | **Accession number** | **UC, average LFQ($10^7$)<sup>a</sup>** | **NBI, average LFQ($10^7$)<sup>a</sup>** | **Fold change UC/NBI<sup>b</sup>** |
| --- | --- | --- | --- | --- |
| Apolipoprotein A-I | APOA1_HUMAN | 249 ± 21 | 57 ± 5.5 | 4.37* |
| Apolipoprotein A-II | APOA2_HUMAN | 2.8 ± 0.4 | 0.1 ± 0.0 | 18* |
| Apolipoprotein A-IV | APOA4_HUMAN | 6.9 ± 1.5 | ND | - |
| Apolipoprotein B-100 | APOB_HUMAN | 628 ± 14 | 64 ± 8.2 | 9.8* |
| Apolipoprotein C-I | APOC1_HUMAN | 0.6 ± 0.0 | ND | - |
| Apolipoprotein C-II | APOC2_HUMAN | 0.1 ± 0.0 | ND | - |
| Apolipoprotein C-III | APOC3_HUMAN | 4.0 ± 0.3 | ND | - |
| Apolipoprotein D | APOD_HUMAN | 4.7 ± 0.7 | ND | - |
| Apolipoprotein E | APOE_HUMAN | 7.0 ± 0.7 | ND | - |
| Apolipoprotein L1 | APOL1_HUMAN | 0.4 ± 0.0 | ND | - |
| Apolipoprotein M | APOLM_HUMAN | 0.1 ± 0.0 | ND | - |
| Apolipoprotein(a) | APOA_HUMAN | 0.5 ± 0.1 | ND | - |

(a) Label-free quantification (LFQ) based on average normalized peak intensity ± SEM of samples isolated by UC (3: #1UC, #2UC, #3UC) or NBI (3: #1NBI, #2NBI, #3NBI); (b) fold-changes in expression between the samples isolated by UC and NBI; \*, $p < 0.05$ , by Wilcoxon-Mann-Whitney test. ND, not determined, proteins not found in NBI samples.

We conclude that NBI is a fast, high-yield procedure to isolate polydisperse circulating EVs that are almost lipoprotein-free with preserved biochemical and biophysical properties. Therefore, we implemented this procedure to isolate EVs from the plasma of our ALS patients and controls to identify possible biomarkers of disease.

### The peculiar size distribution of polydisperse plasma EVs in ALS patients

Using the purification protocol with NBI we isolated EVs from the plasma of 106 ALS patients and 96 controls (Table 1), 36 healthy controls (HC) and 60 disease controls: 28 patients with MD that generally do not imply nerve damage, and 32 patients with SBMA, an ALS-mimic motor neuron disease. First, we investigated whether ALS patients could be distinguished from controls on the basis of the EV concentration and diameter. We calculated the zeta potential of EVs, the measure of the net charge of EV membranes and transmembrane proteins, and found no significant differences among the groups (values between -7.8 and -9.8 mV; Supplementary Fig. 3a). To detect the EV number and size distribution, we did a nanotracking analysis (Nanosight, Malvern). We analyzed the size distribution of plasma EVs within the 50-300 nm range in ALS patients and controls (Fig. 2a). HC presented two peaks (blue line) that were not preserved in all three diseases (ALS, MD, and SBMA). The average curve of the SBMA patients had a higher peak in the distribution (green line), while ALS and MD showed similar distribution (red and yellow line, respectively).

**Figure 2.**
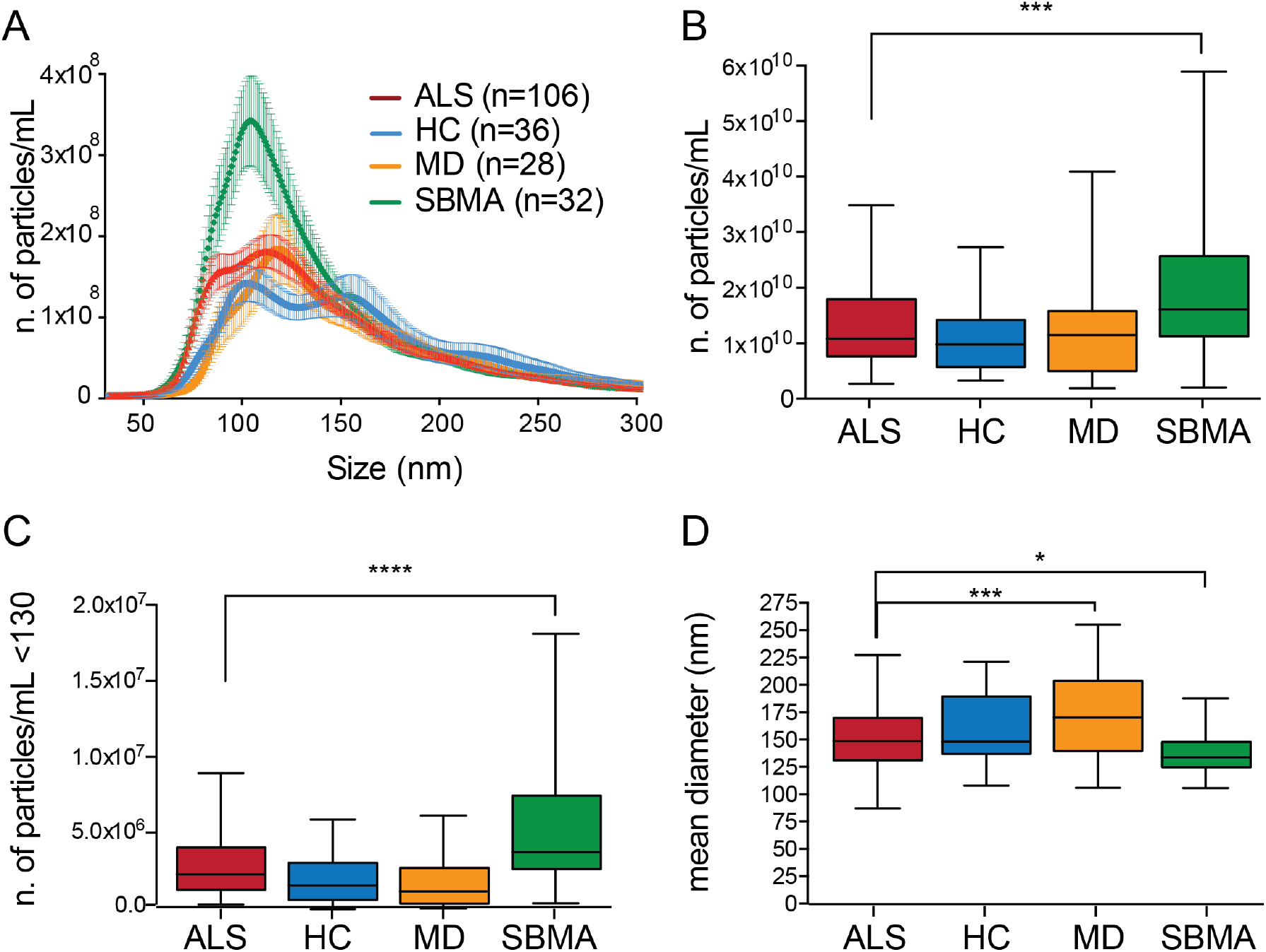
ALS plasma EVs are differentially distributed in size and amount compared to HC, MD and SBMA plasma EVs. **a** Representative average curve of size distribution of ALS, HC, MD and SBMA. **b** Box-plots showing the number of particles per mL (no. particles/mL) of plasma EV in the four groups; ALS, HC, MD, SBMA; one-way ANOVA, p=0.0006; ***p=0.0004 for ALS versus SBMA by Dunnett’s multiple comparisons test. **c** Box-plots showing the number of particles per mL below 130 nm; one-way ANOVA, p<0.0001; ****p<0.0001 for ALS versus SBMA by Dunnett’s multiple comparisons test. **d** Box-plots showing the mean diameter (nm) of EVs in the four groups; one-way ANOVA, p<0.0001; ***p=0.0005 for ALS versus MD and *p=0.037 for ALS versus SBMA by Dunnett’s multiple comparisons test. **b-d** Only significant pairwise comparisons, ALS versus HC/DM/SBMA, were indicated. ALS, amyotrophic lateral sclerosis plasma EVs; HC, healthy control plasma EVs; MD, muscle dystrophy plasma EVs; SBMA, spinal and bulbar muscular atrophy plasma EVs.

Looking at the total amount of particles per mL, we saw a significant difference in EV number between the two motor neuron diseases, ALS and SBMA, with an average between 1.1E+10 and 2E+10 particles/mL (Fig. 2b). We wondered whether the number of small vesicles could distinguish the different groups. We considered the median of all the samples as the cut-off (130 nm) between the smaller and bigger populations. The number of particles/mL below the cut-off differed significantly in the four groups, with the highest number of EVs in samples from ALS and SBMA (2.7E+04 ± 2.1E+03 and 5.4E+04 ± 7.5E+03, respectively) (Fig. 2c).

Finally, we compared the sizes of EVs and, surprisingly, MD EVs had the largest mean diameter (174 ± 7 nm) differing significantly from ALS EVs and SBMA EVs (mean diameters respectively 151 ± 3 nm and 137 ± 3 nm) (Fig. 2d). D50 was also higher in MD than ALS and SBMA (Supplementary Fig. 3d). ALS mean diameter and D50 were significantly larger than SBMA EVs, so SBMA EVs were the smallest of these different conditions (Fig. 2d, Supplementary Fig. 3d). The mode diameter, D10 and D90 presented a similar pattern (Supplementary Fig. 3b,c,e).

### The distinctive size distribution of polydisperse plasma EVs in two ALS mouse models

To test whether the differences in size distribution in ALS plasma EVs were recapitulated in experimental models, we analyzed the plasma-derived EVs purified from two transgenic ALS mouse models expressing either SOD1^G93A^ [21] or TDP-43^Q331K^ [22]. These two mouse models of motor neuron disease are pathologically very different, in particular for their rate of disease progression, which is fast and slow respectively, and offer the opportunity to study plasma EVs independently from genotype and phenotype. SOD1^G93A^ mice show early symptoms at 10 weeks of age, and become symptomatic with loss of body weight, muscular weakness, and motor impairment at 16 weeks. TDP-43^Q331K^ mice present 30% loss of L5 motor axons and 30-45% loss of lower motor neurons by 10 months of age. We analyzed the size distribution of particles in control and transgenic mice at different ages (Fig. 3a and e). The peak of the plot was substantially higher and shifted toward smaller size. In fact, the mean number of particles was significantly larger (1.7 times) in symptomatic mice expressing either SOD1^G93A^ or TDP-43^Q331K^, suggesting similar changes in EV production or in the rate of elimination in both genetic models (Fig. 3b and f). There was already a tendency to an increase in particle number at a presymptomatic stage of the disease (Fig. 3b). This difference became significant when we considered the number of particles/mL below the 130 nm cut-off for the TDP-43^Q331K^ mice (Fig. 3c and g). As in the sporadic patients, the mean diameter was also smaller in both ALS mouse models, 11% less in symptomatic SOD1^G93A^ mice and 14% in the TDP-43^Q331K^ mice (Fig. 3d and h), further indicating a possible association of this parameter with motor neuron degeneration and denervation.

**Figure 3.**
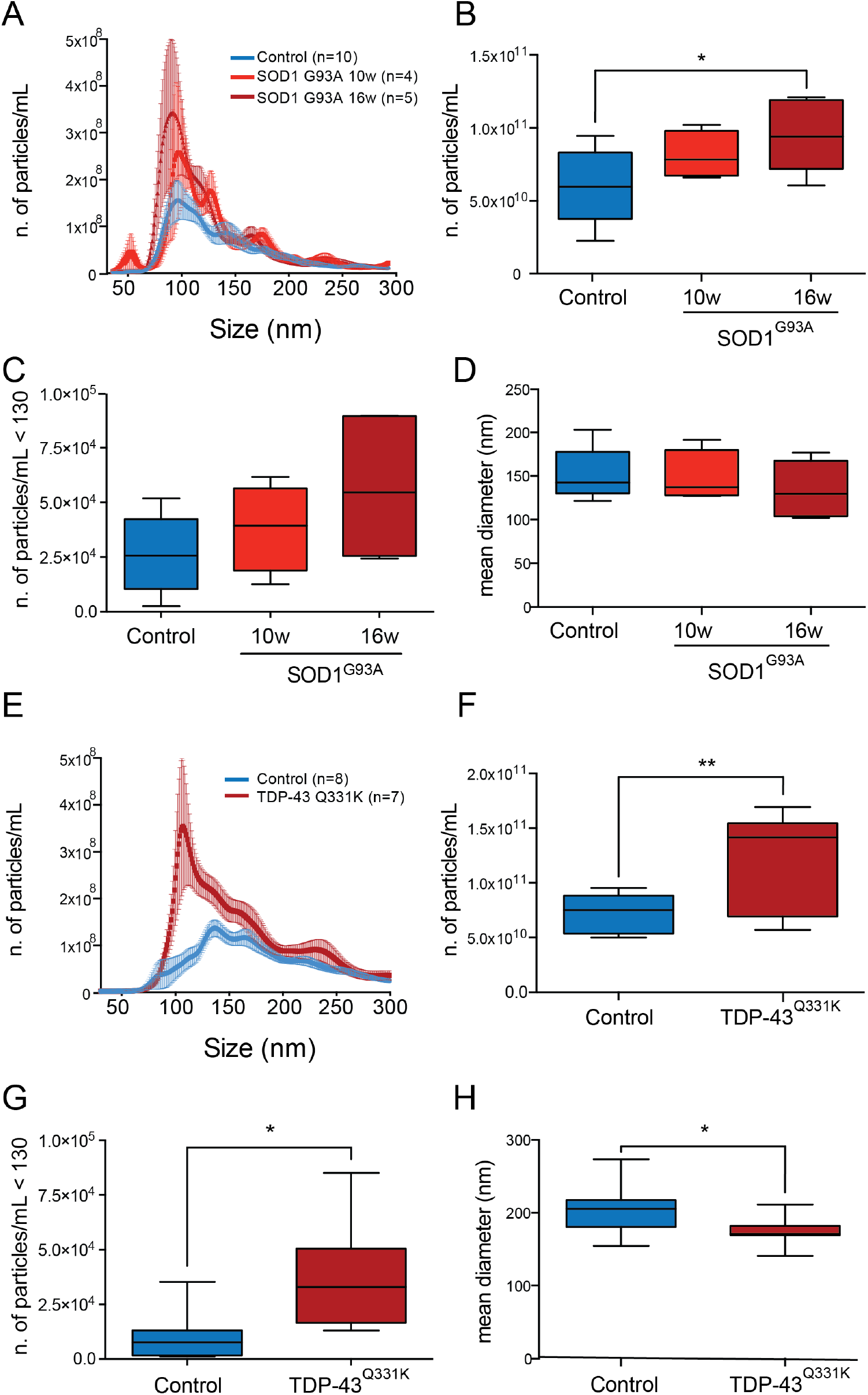
Mouse plasma EVs from two ALS mouse models have peculiar size distribution, similarly to human ALS plasma EVs. **a, e** Representative average curve of size distribution of SOD1^G93A^ mice at 10 and 16 weeks and of TDP-43^Q331K^ mice at 10 months and age-matched controls. **b** Box-plot showing the number of particles per mL in plasma of SOD1^G93A^ mice at 10 and 16 weeks, compared to controls. One-way ANOVA, p=0.029; *p=0.028 between SOD1^G93A^ 16 weeks and controls by Tukey’s multiple comparisons test. **c** Box-plots showing the number of particles per mL below 130 nm of SOD1^G93A^ EVs at 10 and 16 weeks compared to controls; one-way ANOVA, p=0.069. **d** Box-plot showing the mean diameter of plasma EVs of SOD1^G93A^ EVs at 10 and 16 weeks compared to controls. One-way ANOVA, p=0.57. **f** Box-plot showing the number of particles per mL in plasma of TDP-43^Q331K^ mice at 10 months compared to controls. Student t-test, **p=0.0063. **g** Box-plots showing the number of particles per mL below 130 nm of TDP-43^Q331K^ mice at 10 months compared to controls. Student t-test, *p=0.012. **h** Box-plot showing the mean diameter of plasma EVs of TDP-43^Q331K^ EVs at 10 months compared to controls. Student t-test, *p=0.039.

These results suggest that the size distribution and the mean size of ALS EVs in two *in vivo* models of the pathology and in patients are parameters that can be used to distinguish ALS from other conditions, possibly already at an early stage.

### Analysis of a panel of EV-associated proteins in ALS patients and mouse models

To determine whether specific proteins, previously associated with ALS, are differentially enriched in plasma-derived EVs of ALS patients compared to the other groups, we set up immunoblotting for two candidate proteins: human and mouse HSP90 and PPIA (Fig. 4a and b; Supplementary Fig. 4).

**Figure 4.**
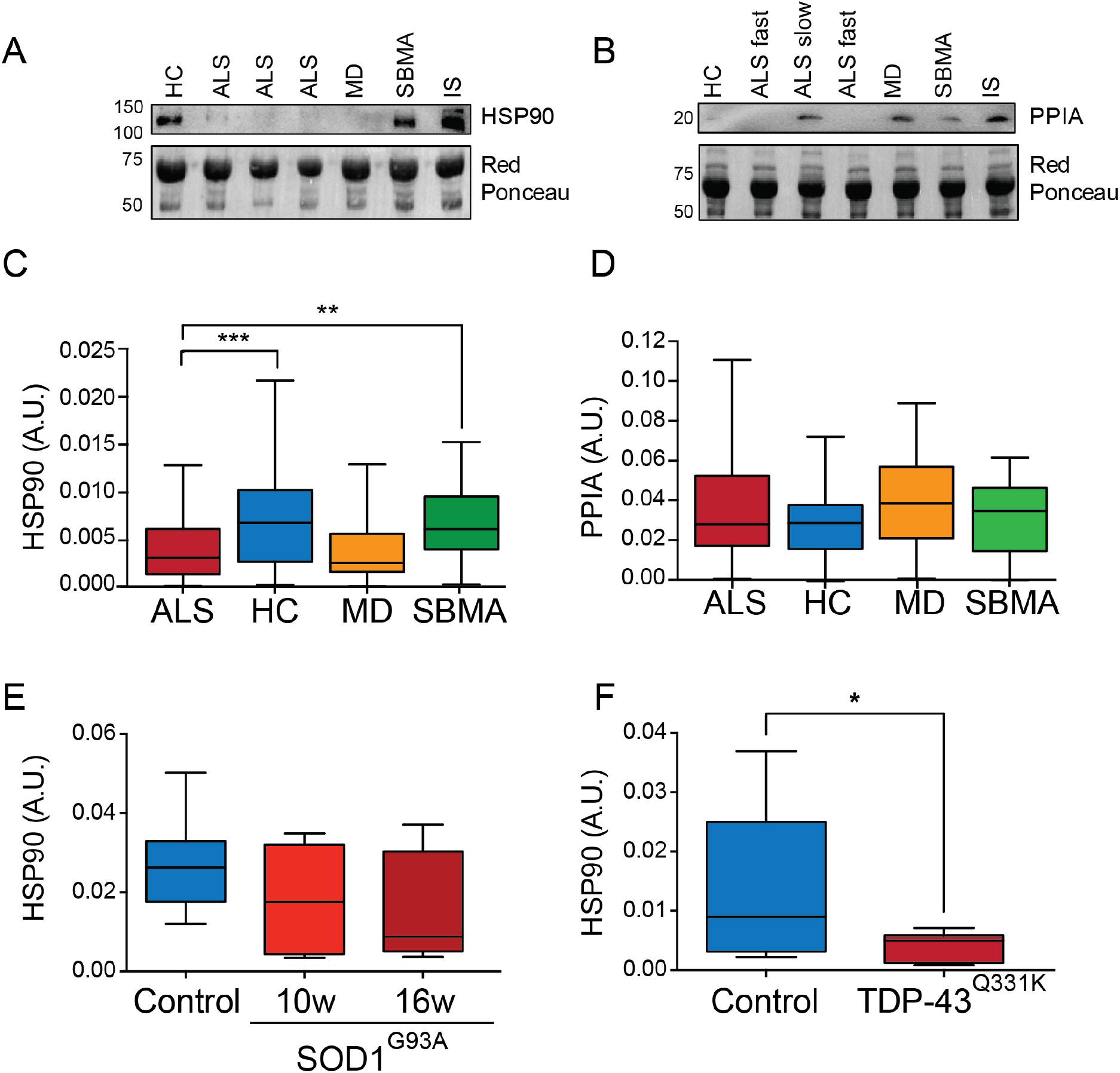
HSP90, but not PPIA, and phosphorylated TDP-43 shows specific enrichment in relation to the disease in human and mouse plasma EVs. **a, b** Representative immunoblotting for HSP-90 (**a**), and PPIA (**b**) and relative Red Ponceau in human EV samples. IS means internal standard. **c** Box-plot showing the levels of HSP90 in the plasma EVs of ALS, HC, MD and SBMA. One-way ANOVA, p<0.0001; ***p=0.0003 ALS versus HC and **p=0.0047 ALS versus SBMA by Dunnett’s multiple comparisons test. **d** Box-plot showing the levels of PPIA in the plasma EVs of ALS, HC, MD and SBMA. one-way ANOVA, p=0.22. **e** Box-plot showing the levels of HSP90 in SOD1^G93A^ plasma EVs at 10 and 16 weeks of age compared to controls. One-way ANOVA, p=0.29. **f** Box-plot showing the levels of HSP90 in TDP-43^Q331K^ plasma EVs at 10 months of age compared to controls. Student t-test, *p=0.037.

HSP90 is a highly abundant, ubiquitous molecular chaperone that supports protein folding. In ALS it has been detected in protein inclusions in SOD1^G93A^ mice and sporadic patients [44], with a consequent decrease in the soluble fraction; it was also unusually low in ALS patients and mouse models with early disease onset and a severe phenotype [45]. It is present in EVs, but its levels have never been analyzed in plasma-derived EVs of ALS patients. HSP90 is significantly less present (52% lower) in the plasma-derived EVs of ALS patients compared to HC and SBMA (Fig. 4c). HSP90 is also low in plasma-derived EVs of symptomatic SOD1^G93A^ (41% less) and TDP-43^Q331K^ mice (73% less) (Fig. 4e-f), suggesting that low levels of HSP90 in plasma EVs could characterize several forms of ALS, sporadic and genetic. Future studies in human genetic forms are now required to confirm this association.

We looked at PPIA, also known as cyclophilin A, a highly abundant, ubiquitous foldase and chaperone involved in TDP-43 trafficking and function [46]. We have previously shown that PPIA is enriched in protein inclusions in SOD1^G93A^ mice and patients [44]. Soluble PPIA is particularly low in ALS patients compared to healthy individuals and subjects with other neurological diseases [47]. However, in contrast with HSP90, PPIA levels did not change among groups, indicating that EV-enriched PPIA may not be useful for differential diagnosis of ALS from other diseases (Fig. 4d).

Finally, to characterize our patients’ cohorts better, we measured phosphorylated neurofilament heavy chain (pNFH) in plasma. As expected, pNFH was substantially higher in ALS than in all other experimental groups, especially fast-progressing patients (Fig. 5a and Fig. 7f). We attempted to measure pNFH in EVs, but we could not detect a reliable signal (data not shown), suggesting that pNFH is probably not loaded in EVs or only at very low concentrations.

**Figure 5.**
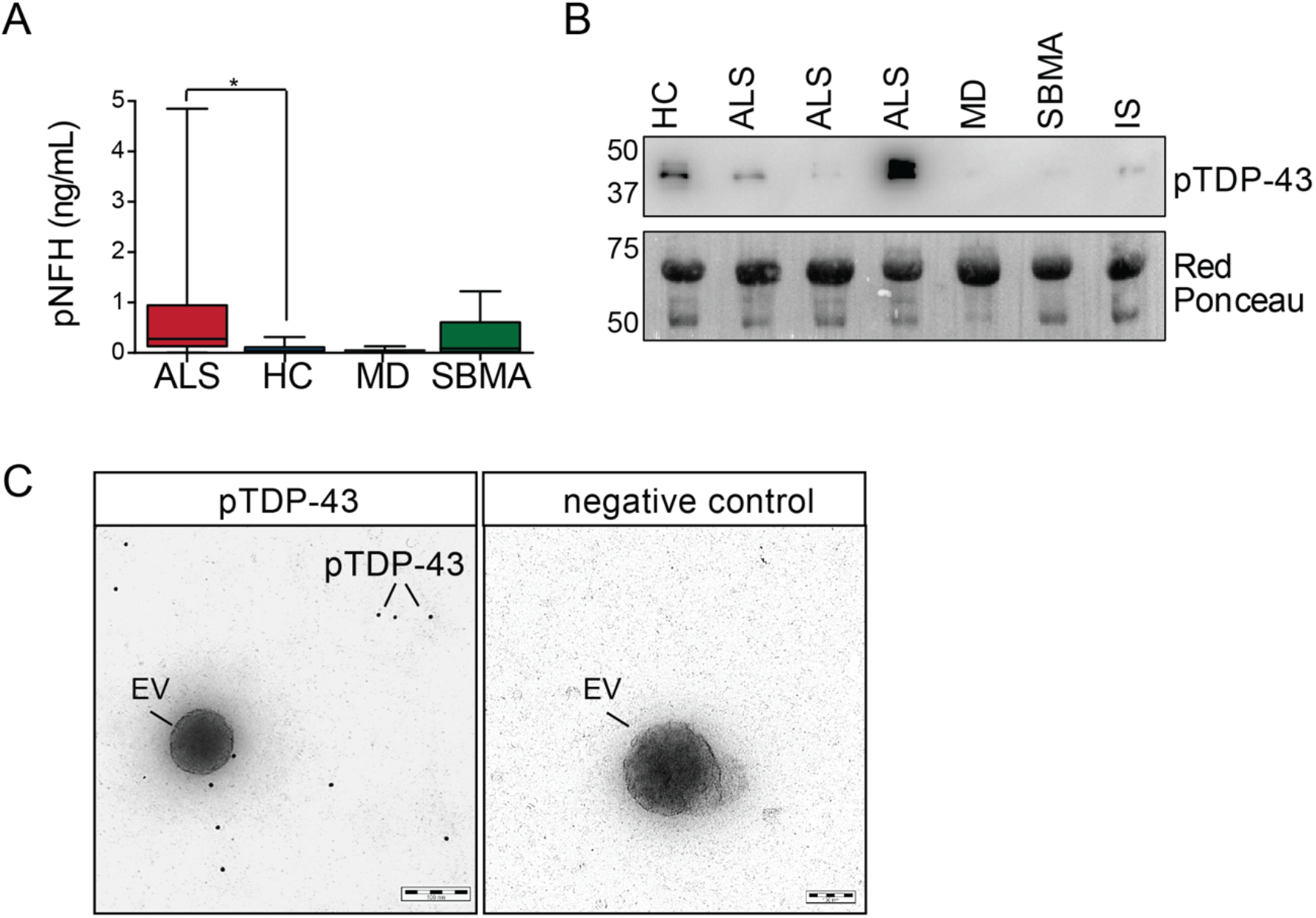
Neurofilaments and phosphorylated TDP-43 are not EV cargos. **a** Plasma levels of phosphorylated neurofilament H (pNFH) in ALS, HC, MD and SBMA. One-way ANOVA, p=0.033; *p=0.043 ALS versus HC by Dunnett’s multiple comparisons test. **b** Representative immunoblotting for pTDP-43 and relative Red Ponceau in human EV samples. IS means internal standard. **c** Immunogold transmission electron microscopy (TEM) analysis of plasma EVs purified with NBI and stained with phosphorylated TDP-43 antibody (pTDP-43) (left panel) or negative control (right panel). Phosphorylated TDP-43 is indicated with a line (12 nm gold nanoparticles).

**Figure 6.**
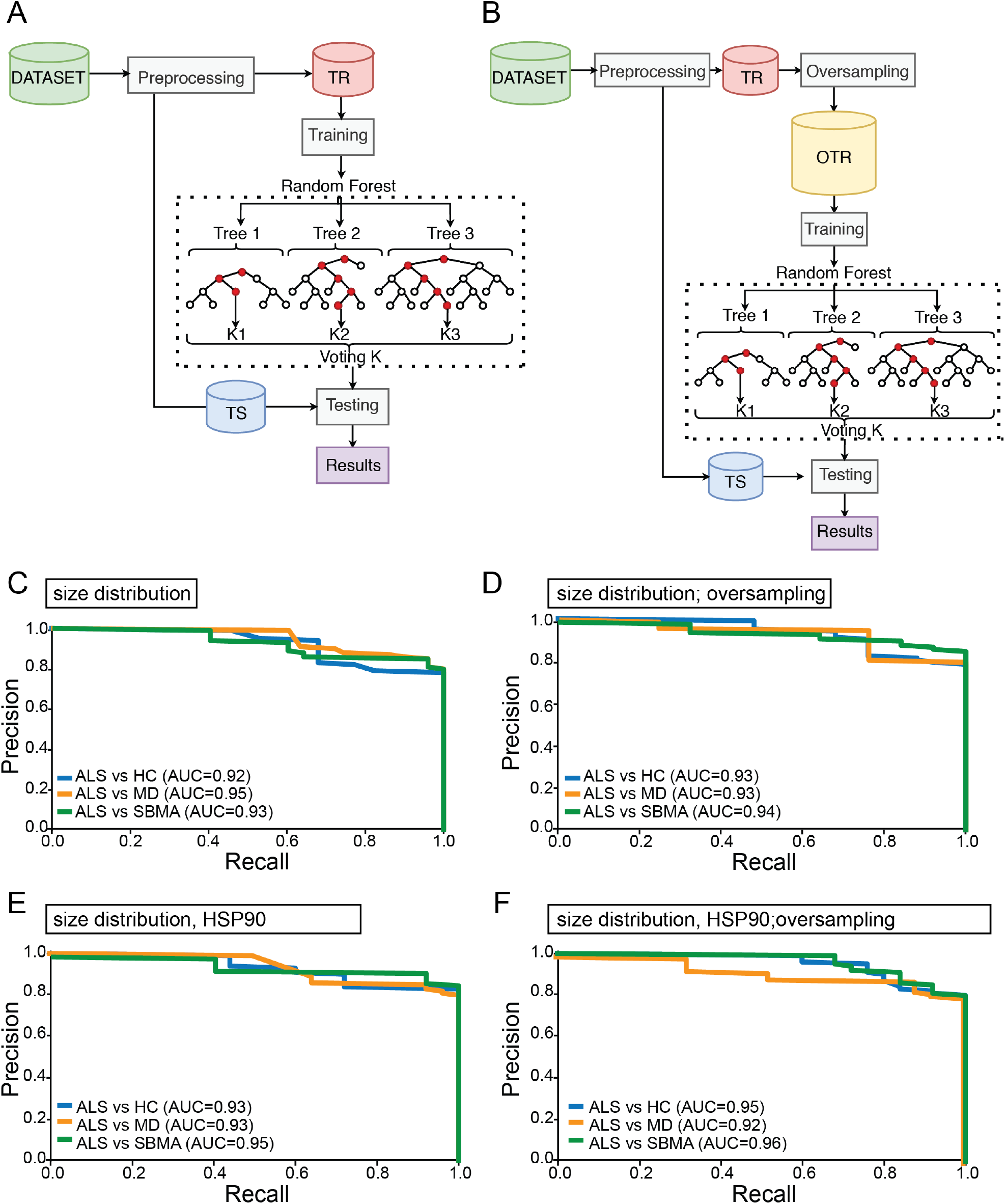
The size distribution and the amount of HSP90 in EVs help to correctly classify ALS patients. **a-b** machines learning frameworks used in the experiments. Scheme **a** is related to the basic framework, while scheme **b** is related to the advanced one. They both consist of three main steps: data handling, training of the models and testing of the models on an independent cohort. Those schemes differ in the data handling phase. The basic framework only preprocesses data, while the advanced one uses a tailored oversampling strategy to handle unbalanced data. TR is training set; TS is test set. **c** Precision-recall curves on size distribution for a binary comparison. **d** Precision-recall curves on oversampled data on size distribution for a binary comparison. **e** Precision-recall curves on size distribution and HSP-90 for a binary comparison. **f** Precision-recall curves on oversampled data on size distribution and HSP-90 for a binary comparison.

**Figure 7.**
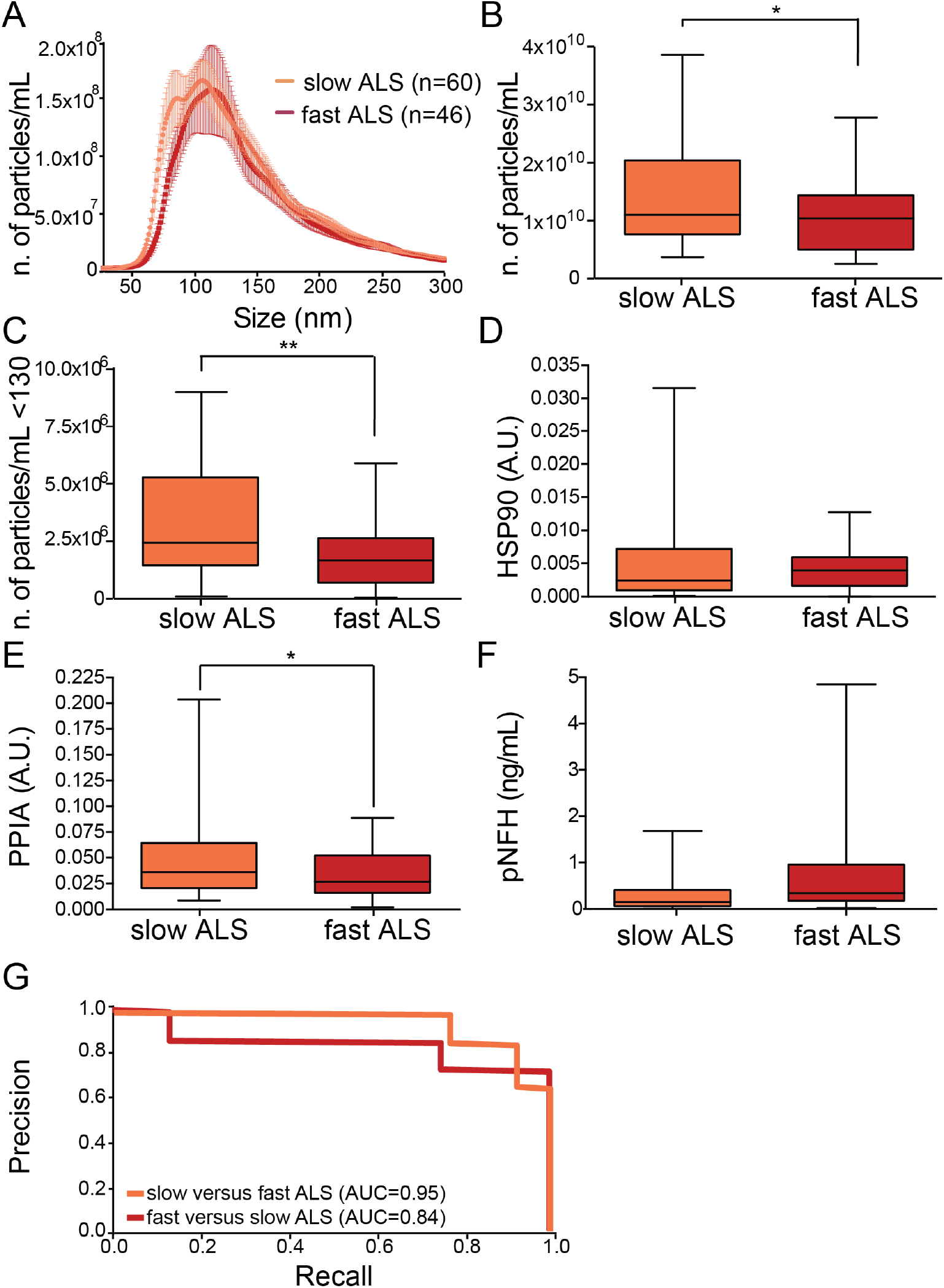
EV parameters distinguish fast and slow-ALS progressors. **a** Representative average curve of size distribution for slow and fast ALS. **b** Numbers of particles per mL in slow and fast ALS; Student t-test, *p=0.017. **c** Number of particles per mL below 130nm in slow and fast ALS; Student t-test, **p=0.0053. **d** Levels of HSP90 in slow and fast ALS; Student t-test, p=0.46. A.U.: arbitrary units. **e** Levels of PPIA in slow and fast ALS; Student t-test, *p=0.015. A.U.: arbitrary units. **f** Plasma levels of pNFH in slow and fast ALS; Student’s t-test, p=0.09. **g** Precision-recall curves on stratified data for ALS progression showing the AUC for each comparison.

We also investigated whether TDP-43, the hallmark of ALS, is detectable in plasma-derived EVs. Previous reports showed insoluble TDP-43 in plasma EVs isolated by UC from ALS patients [17]. Using our approach, we observed a small amount of TDP-43 in human plasma-derived EVs, as indicated by two different antibodies directed toward the N or the C terminus of the protein, strongly cross-reacting in these conditions with plasma protein contaminants, immunoglobulins, and albumin (Supplementary Fig. 5a and b). We tested whether the anti-phosphorylated TDP-43 antibody was a better tool to analyze TDP-43 in EVs. Surprisingly, the antibody revealed a specific and distinct doublet at 45 kDa (Supplementary Fig. 5c-e). To prove that phosphorylated TDP-43 is contained in the EVs, we performed immunogold TEM in three independent ALS samples. Unexpectedly, no signal for phosphorylated TDP-43 was revealed inside the EVs, suggesting that phosphorylated TDP-43 is not an intravesicular cargo in recovered plasma EVs (Fig. 5c).

We conclude that among our candidate protein markers, only HSP90 is differentially represented in ALS EVs than other diseases and can be used in a panel of markers to define the fingerprint of ALS plasma-derived EVs.

### The ability of EV parameters alone and in combinations to distinguish ALS from MD, SBMA and healthy controls

To test whether the parameters we analyzed can be used to distinguish ALS patients from other groups, we employed machine learning techniques. First, we tested whether the size distribution of plasma EVs can distinguish ALS patients from others. As a preliminary step, we normalized all the distribution curves (Supplementary Fig. 3f). Then we used machine learning tools to compress the signal [30,31] (Supplementary Fig. 3g), and used the compressed data to build a machine learning model based on Random Forests (RF) (Fig. 6a). We evaluated the model’s ability to distinguish ALS patients from the others (HC, MD, and SBMA). The Precision-recall curves provided the AUC, ranging between 0.92 and 0.95 (Fig. 6c). We oversampled our datasets with MWMOTE (Fig. 6b) [32] and the prediction increased to between 0.93 and 0.94 (Fig. 6d).

We wondered whether combining the size distribution parameters with the values obtained for HSP90 in the EVs would improve the disease classification. The new models showed an AUC of 0.93 for ALS versus HC, 0.93 for ALS compared to MD, and 0.95 for ALS compared to SBMA. The oversampling only enhanced the classification of ALS when tested with HC (AUC=0.93 vs. 0.95) and with SBMA (AUC=0.95 vs 0.96) (Fig. 6f).

The machine learning approach indicates that the size distribution of EVs can be used to classify patients and controls, with relatively good prediction. Oversampling slightly improved the AUC in almost all comparisons. The models combining the size distribution and HSP90 reached excellent AUC values, particularly for distinguishing ALS from SBMA.

### Specific parameters like the number of particles, size, and PPIA distinguish fast from slow patients

We wondered whether the parameters we analyzed could distinguish slow from fast progressors. We found no significant correlation between EV parameters and clinical variables (ALSFRS-R, disease duration, site of onset, ALSFRS-R) by univariable and multivariable linear models. However, ALS patients stratified according to disease progression (slow- and fast-ALS) showed a shift in the peaks of the plot of the average size distribution (Fig. 7a).

We stratified the data on EV particles/mL, the particles smaller than 130 nm, HSP90, and PPIA. Slow-ALS patients had a significant, 1.3-fold increase in EV particles/mL compared to fast-ALS, indicating that average EV concentration might be a selective biomarker of slow-ALS (Fig. 7b). This was confirmed for particles smaller than 130 nm (1.6-fold increase) (Fig. 7c). We then decided to test whether HSP90, and PPIA were differentially enriched in slow-ALS and fast-ALS plasma-derived EVs. HSP90 did not show any difference, suggesting that its protein level is useful for distinguishing ALS from other diseases, but does not change with the rate of progression (Fig. 7d).

While PPIA did not change across different diseases (Fig. 4d), it was 32% lower in fast-ALS than slow-ALS, confirming our previous finding that patients with a smaller amount of PPIA present earlier onset and faster progression of the disease [45,47] (Fig. 7e). Interestingly, fast-progressing patients had high pNFH plasma levels, though not significantly different from slow-progressing patients, indicating that at least in our patients, pNFH cannot distinguish between fast and slow progression (Fig. 7f).

We built a new mathematical model with the stratified data according to the rate of ALS progression, for size distribution, and PPIA. When we oversampled the data the AUC for fast and slow ALS ranged between 0.84 and 0.95, suggesting that these three parameters could be exploited for ALS prognosis (Fig. 7g).

## Discussion

Several approaches have been reported to successfully purify EVs from plasma, like ultracentrifugation, filtration, precipitation, chromatography, immunocapture, microfluidics, but there is still no consensus on which of these techniques give the best results in terms of yield, purity and physical integrity [48]. In the clinical setting, fast, reproducible, easy protocols are needed. Here we showed that NBI, a recently established purification method requiring fast, simple procedures, allows the recovery of a larger number of plasma-derived EVs. Therefore, NBI is suitable for large and complex clinical studies in which multiple outcomes are measured. In fact, few hundreds µL of plasma per subject are enough to perform an array of downstream analyses for EV characterization.

Plasma EVs are contaminated by the abundant plasma proteins, especially lipoproteins (HDL, LDL, VLDL, and chylomicrons), which are much more abundant than EVs and biophysically very similar in terms of density and size [43], substantially affecting downstream analysis. Combined isolation approaches are generally used to ensure the purity of EVs, but the continuous sample manipulation may affect the EV integrity and their biophysical features [49]. Here we report that EVs purified with NBI had minimal lipoprotein contamination, offering a new strategy for plasma EV purification and analysis. EV-like liposomes also retained their mean and mode diameter when isolated with NBI. It is probably essential to use a technique that preserves EV integrity, minimizing EV aggregation [20]. This observation stresses the differences in mean diameter, mode, D10, D50, and D90 between ALS and MD samples.

We did not examine different plasma-EV subpopulations, so we can only speculate that different proportions of EV subclasses, deriving from different organs or cell populations, may be present in ALS samples, compared to the other conditions, resulting in a smaller mean diameter and more particles in human and mouse plasma EVs. Furthermore, the actual EV size depends not only on the type of membrane phospholipid but also on the presence or absence of particular membrane proteins [50]. EVs vary widely in molecular composition, and their surface proteins bear characteristics of their tissues of origin [51], suggesting that in each pathological condition plasma EVs released from the damaged tissue may have a specific size distribution. For example, brain-derived EVs in plasma have already been reported in Alzheimer’s disease patients up to ten years before the onset of the disease [52], although there is no specific characterization of their mean size yet. Similarly, muscle-derived EVs have been studied for their contribution to MD [53], but no direct comparison between brain and muscle in degenerative conditions has been made yet in terms of their specific contribution to plasma EVs. Here we show that the EVs in ALS differ substantially from MD patients. SBMA is a neuromuscular disease with early muscle degeneration and motor neuron loss. It presents EV features that are similar to ALS EVs but are not wholly different from the MD.

Another important aspect of EVs is the proteomic and genomic material in the lumen. EVs have been considered means of intercellular disease spread both in cancer and in neurodegenerative diseases, where they contribute to the formation of metastasis and the seeding of protein inclusions, respectively [54,55]. Proteins directly involved in ALS, like TDP-43, SOD1, FUS and the dipeptide repeat proteins derived from C9orf72 aberrant repeats were detected in EVs from *in vitro* and *in vivo* models, but no functional role in the disease spread *in vivo* has been provided yet [15,16,56–59]. Nevertheless, insoluble mutant proteins were observed in plasma-derived microvesicles of ALS patients, suggesting that there may be ALS-associated proteins in plasma-derived EVs [17]. Here we report a clear-cut signal for phosphorylated TDP-43 in plasma EVs using a phosphorylation-dependent antibody that stains pathological ubiquitin-positive inclusions in ALS and frontotemporal lobar degeneration (FTLD) patients [61]. In our analysis the signal for the phosphoprotein was high even when TDP-43 was hardly detectable by two pan antibodies, suggesting that there was a significant enrichment of hyperphosphorylated TDP-43 in EVs isolated from plasma. However, phosphorylated TDP-43 was not observed inside the EVs by TEM therefore cannot be considered an intravesicular cargo. We can assume that phosphorylated TDP-43 may have a certain affinity for the EV “protein corona”, which comprises a variety of plasma proteins such as apolipoproteins, immunoglobulins, complement factors and coagulation factors that are bound to EV membranes as much as to any nanomaterials in contact with biofluids [62,63]. This phenomenon has been clearly observed independently from the EV isolation method. Although EV-associated TDP-43 does not distinguish ALS patients from healthy controls and cannot be considered a useful marker, it may be interesting to study the effect of phosphorylated TDP-43 on EV external membrane for the EV cellular uptake and pathophysiological role. Furthermore, whether TDP-43, phosphorylated or not, is an intra-or an extravesicular cargo in EVs isolated from CSF has not been demonstrated yet along with a robust method for TDP-43 quantification in biofluids [4,64].

HSP90, a critical molecular chaperone of the protein quality control complex [65], is significantly lower in ALS EVs in patients and two genetic ALS mouse models. This agrees with our previous findings, that HSP90 is entrapped in insoluble proteins in the spinal cord of sporadic patients and late symptomatic SOD1^G93A^ mice [44]. We also observed low levels of HSP90 in SOD1^G93A^ astrocytes [16] and PBMCs, and in ALS sporadic patients [45], suggesting that HSP90 could be measured in plasma-derived EVs to distinguish ALS patients from other conditions involving motor neurons, like SBMA.

The levels of PPIA did not change across diseases, but there were significantly lower in fast-ALS than slow progressors. PPIA, also known as cyclophilin A, is a multifunctional protein with foldase and molecular chaperone activities and is one of the most commonly identified proteins in EVs [66]. PPIA is also an interacting partner of TDP-43 and regulates its trafficking and function [46]. PPIA is significantly enriched in the insoluble fraction of spinal cords of ALS patients and mice [44], and its soluble form is reduced in PBMC of sporadic ALS patients [45]. Therefore, it is not surprising that in fast-ALS, a severe condition associated with diffuse TDP-43 pathology, PPIA is less present in EVs, possibly stacked intracellularly while contrasting protein aggregation. We recently reported that low PPIA soluble levels in ALS PBMC were associated with six months earlier death [47], indicating that EVs reflect pathological intracellular alterations and supporting plasma-derived EVs as promising predictors of ALS disease progression.

Neurofilaments are generally increased in acute and chronic neurodegenerative conditions [10]. They are released into the biofluids as the final event that leads to the degeneration of motor neurons. Their levels correlate well with the severity of the axonal degeneration, presenting a smaller increase in motor neuron diseases with a milder phenotype, like SBMA [67]. Despite their lack of specificity, they are considered the best biomarkers to diagnose and predict ALS progression [68,69]. We measured pNFH in the plasma of all our samples and confirmed the significant higher levels in ALS compared to all the other diseases and HC, confirming its value as a diagnostic biomarker. However, when we stratified our samples, EV concentration and size, PPIA, HSP90, and phosphorylated TDP-43 significantly distinguished fast- and slow-ALS, but pNFH did not. There was only a trend towards higher pNFH in fast ALS, indicating that plasma pNFH is not a robust prognostic biomarker. This was confirmed very recently in a large longitudinal study in which, in contrast with neurofilament light, pNFH has shown little prognostic value [70]. We did not detect neurofilaments in EVs, suggesting that neurofilaments mainly reflect axonal damage and are released freely in the biofluids.

Machine learning models offer unprecedented opportunities to evaluate the potential of specific targets as biomarkers, even with considerable sample size limitations. Machine learning techniques have already been applied to ALS clinical and imaging data sets and resulted in promising diagnosis and prognosis models [71]. They have been exploited for ALS progression and survival ranking [72–74], for predicting the outcome of specific treatments (e.g., riluzole) based on the patient’s characteristics [75], and the use of candidate protein amounts in the CSF as pharmacodynamic biomarkers [76,77]. Here we describe the first model built on EV biophysical parameters that offers great potential for differential diagnosis of ALS compared to MD and SBMA. We also report that including the amount of plasma EVs HSP90 and PPIA in the mathematical model increases its predictive power.

## Conclusions

ALS is clinically, genetically, and neuropathologically highly heterogeneous and biomarkers for diagnosis, prognosis and stratification are lacking [78]. This makes it difficult to develop effective therapies. We set up a biomarker analysis in plasma EVs that is at the same time methodologically robust and easy-to-perform, which makes the translation to the clinics of our findings straightforward. We show that EV biophysical parameters and protein cargoes are promising means to characterize disease’s complexity. Our mathematical model integrating EV size distribution data with protein cargoes can distinguish ALS from healthy and diseased controls, and classify ALS patients with variable disease progression with high accuracy, confirming the considerable potential of machine learning techniques in improving clinical trial design toward the development of personalized therapies [79]. Validation studies in longitudinal cohorts of patients, starting in proximity of symptom onset are now necessary to verify their applicability in early diagnosis and monitoring treatment efficacy.

## Supporting information

Supplemental Fiigures

## Data Availability

Data availability
All the data generated for this manuscript are available upon requests to the corresponding authors.
Proteomics The whole mass spectrometry proteomics data have been deposited to the ProteomeXchange Consortium (http://proteomecentral.proteomexchange.org/cgi/GetDataset) via the PRIDE partner repository with the data set identifier PXD020629

## Abbreviations

ALS: Amyotrophic lateral sclerosis
AUC: area under the curve
CNS: central nervous system
CSF: cerebrospinal fluid
EVs: Extracellular vesicles
FTLD: Frontotemporal lobar degeneration
HC: Healthy controls
HSP90: Heat shock protein-90
MD: Muscular dystrophies
NTA: Nanotracking analysis
NBI: Nickel-based isolation
PBMCs: peripheral blood mononuclear cells
PPIA: peptidyl-prolyl cis-trans isomerase A
pNFH: phosphorylated neurofilament heavy chain
SBMA: Spinal and bulbar muscle atrophy
SOD1: Superoxide dismutase 1
TEM: Transmission electron microscopy
TRPS: Tunable resistive pulse sensing
UC: Ultracentrifugation
WB: Western blot

## Declarations

### Ethical approval and consent to participate

The study was performed according to the ethical principles of the Declaration of Helsinki and was approved by the local ethics committees. Written informed consent was obtained from all subjects.

### Consent for publication

All authors have given their consent for publication.

### Availability of supporting data

The mass spectrometry proteomics data have been deposited at the ProteomeXchange Consortium (http://proteomecentral.proteomexchange.org/cgi/GetDataset) via the PRIDE partner repository with the data set identifier PXD020629. All other data supporting the present findings are available in the article and its supplemental information files.

### Authors’s contributions

VB and MB were responsible for conception and design of the study. All authors contributed to data acquisition and analysis. VB, MB and LP contributed to drafting the text and preparing the figures. All authors critically evaluated and approved the final manuscript.

## Acknowledgements

We would like to thank Francesca Baldelli Bombelli for the initial discussion on sample analysis, the BioBank of the “Casa di Cura Privata del Policlinico (CCPP)” for providing plasma samples of healthy individuals and Cinzia Bertolin for collecting the SBMA plasma. We thank Judith Baggott for editorial assistance.

## Authors’ information

Manuela Basso and Valentina Bonetto are co-corresponding and co-last authors

## Affiliations

**Istituto di Ricerche Farmacologiche Mario Negri IRCCS, Milano (Italy)**

Laura Pasetto, Laura Brunelli, Giovanna Sestito, Roberta Pastorelli, Elisa Bianchi, Alessandro Corbelli, Fabio Fiordaliso, Valentina Bonetto

**Department of Mathematics “Tullio Levi-Civita”, University of Padova**

Stefano Callegaro, Francesco Rinaldi

**Department of Cellular, Computational and Integrative Biology – CIBIO, University of Trento, Trento, Italy**

Deborah Ferrara, Vito D’Agostino, Alessandro Quattrone, Manuela Basso

**Consiglio Nazionale delle Ricerche, Istituto di Scienze e Tecnologie Chimiche “Giulio Natta” (SCITEC-CNR), Milano, Italy**

Marina Cretich, Marcella Chiari

**Centre for Materials and Microsystems, Fondazione Bruno Kessler, Trento, Italy & Istituto di Biofisica, Consiglio Nazionale delle Ricerche, Trento, Italy**

Cristina Potrich

**‘Rita Levi Montalcini’ Department of Neuroscience, Università degli Studi di Torino, Torino, Italy**

Cristina Moglia, Andrea Calvo, Adriano Chiò

**Department of Neurorehabilitation Sciences, Casa Cura Policlinico (CCP), Milano, Italy**

Massimo Corbo

**Department of Neuroscience, University of Padova, Padova, Italy**

Gianni Sorarù

**NEuroMuscular Omnicentre (NEMO), Serena Onlus Foundation, Milano**

Christian Lunetta

**Department of Neurorehabilitation, ICS Maugeri IRCCS, Milano, Italy**

Gabriele Mora

**Department of Biomedical Sciences (DBS), University of Padova, and Veneto Institute of Molecular Medicine (VIMM), Padova, Italy**

Maria Pennuto

**Corresponding authors**

Correspondence should be addressed to Manuela Basso or Valentina Bonetto

