## Supplemental Fiigures for "Decoding distinctive features of plasma extracellular vesicles in amyotrophic lateral sclerosis"

***Plasma extracellular vesicles and ALS***

Laura Pasetto[1], Stefano Callegaro[2], Alessandro Corbelli[1], Fabio Fiordaliso[1], Deborah Ferrara[3], Laura Brunelli[1], Giovanna Sestito[1], Roberta Pastorelli[1], Elisa Bianchi[1], Marina Cretich[4], Marcella Chiari[4] Cristina Potrich[5], Cristina Moglia[6], Massimo Corbo[7], Gianni Sorarù[8], Christian Lunetta[9], Andrea Calvo[6], Adriano Chiò[6], Gabriele Mora[10], Maria Pennuto[11,12], Alessandro Quattrone[3], Francesco Rinaldi[2], Vito D'Agostino[3], Manuela Basso[1,3*], and Valentina Bonetto[1*]

[1]Istituto di Ricerche Farmacologiche Mario Negri IRCCS, Milano (Italy)

[2] Department of Mathematics "Tullio Levi-Civita", University of Padova

[3]Department of Cellular, Computational and Integrative Biology – CIBIO, University of Trento, Trento, Italy

[4]Consiglio Nazionale delle Ricerche, Istituto di Scienze e Tecnologie Chimiche "Giulio Natta" (SCITEC-CNR), Milano, Italy

[5] Centre for Materials and Microsystems, Fondazione Bruno Kessler, Trento, Italy & Istituto di Biofisica, Consiglio Nazionale delle Ricerche, Trento, Italy

[6]'Rita Levi Montalcini' Department of Neuroscience, Università degli Studi di Torino, Torino, Italy

[7]Department of Neurorehabilitation Sciences, Casa Cura Policlinico (CCP), Milano, Italy

[8] Department of Neuroscience, University of Padova, 35122 Padova, Italy

[9]NEuroMuscular Omnicentre (NEMO), Serena Onlus Foundation, Milano

[10]Department of Neurorehabilitation, ICS Maugeri IRCCS, Milano, Italy

[11] Department of Biomedical Sciences (DBS), University of Padova, 35131 Padova, Italy.

[12] Veneto Institute of Molecular Medicine (VIMM), 35129 Padova, Italy.

*These are co-corresponding and co-last authors.

Supplementary Figure 1

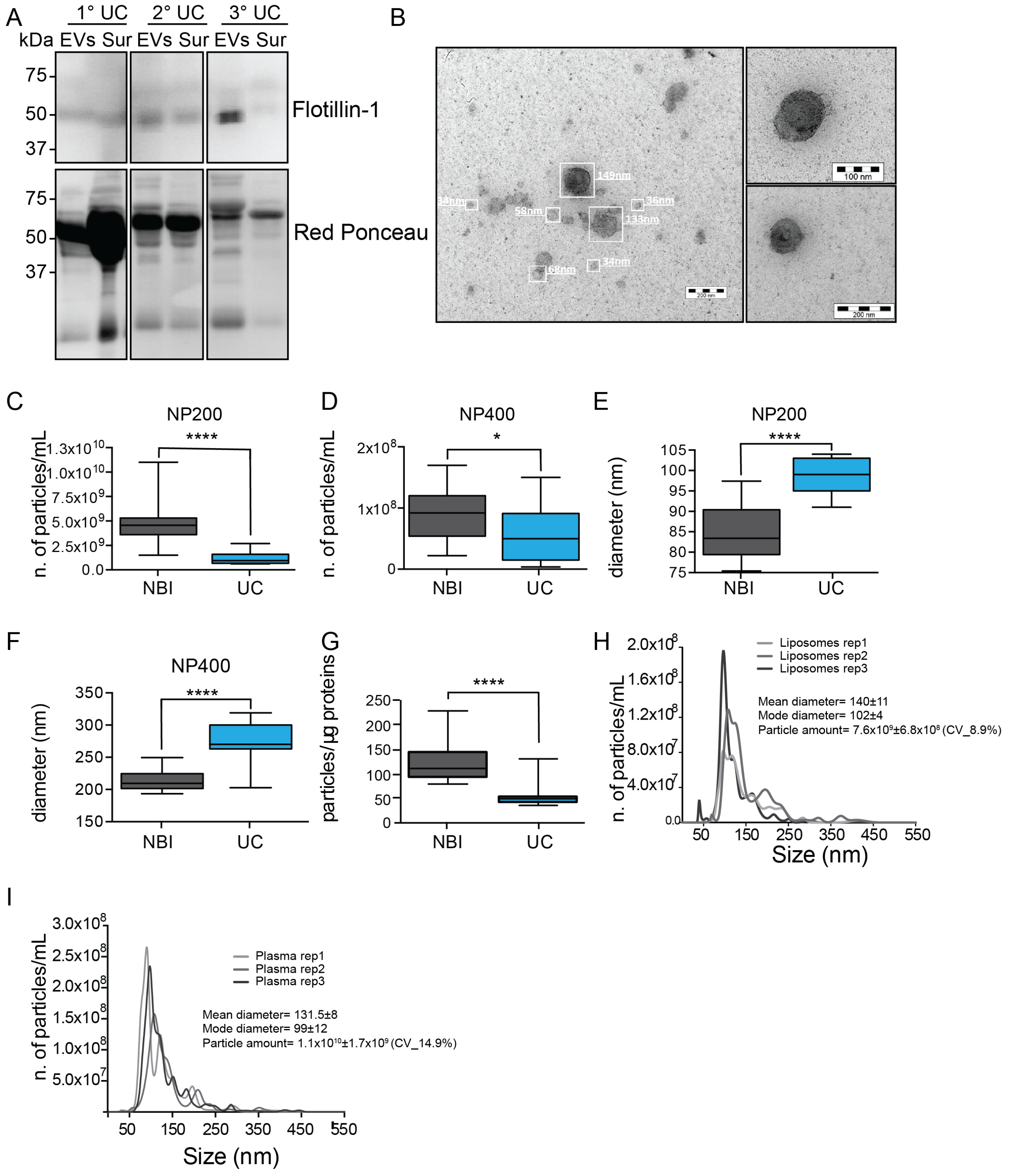


**Supplementary Figure 1.** NBI enriches for a higher number and a smaller average diameter plasma EVs than classical ultracentrifugation (UC). **a** Immunoblotting for flotillin-1 in a pool of human plasma and relative Red Ponceau. ‘1°-2°-3°UC’ stands for first, second, third ultracentrifugation. ‘EVs’ and ‘Sur’ are the pellet and supernatant after the UC. **b** TEM of EVs purified with UC. Bar, 200 nm on the left; 100 nm and 200 nm in the insets on the right. **c** TRPS analysis for the particle amount per mL (n. particles/mL) with the NP200 nanopore of control plasma EVs (n=15) extracted with either NBI or UC. Student t-test; ****p<0.0001. **d** TRPS analysis with the NP400 nanopore of control plasma EVs (n=15) for the particle amount per mL (n. particles/mL) extracted with either NBI or UC. Student t-test; *p=0.037. **e** TRPS analysis with the NP200 nanopore of control plasma EVs (n=15) for the mean diameter (nm) extracted with either NBI or UC. Student t-test; ****p<0.0001. **f** TRPS analysis with the NP400 nanopore of control plasma EVs (n=15) for the mean diameter (nm) extracted with either NBI or UC. Student t-test; ****p<0.0001. **g** Purity index for samples extracted with NBI and UC, calculated by the ratio between the number of particles and the total micrograms of proteins detected in the relative EV samples; Student t-test; ****p<0.0001. **h** Size distribution of liposomes isolated with three independent NBI extractions (Liposomes rep1, 2, 3). **i** Size distribution of plasma isolated with three independent NBI extractions (Plasma rep1, 2, 3).

Supplementary Figure 2

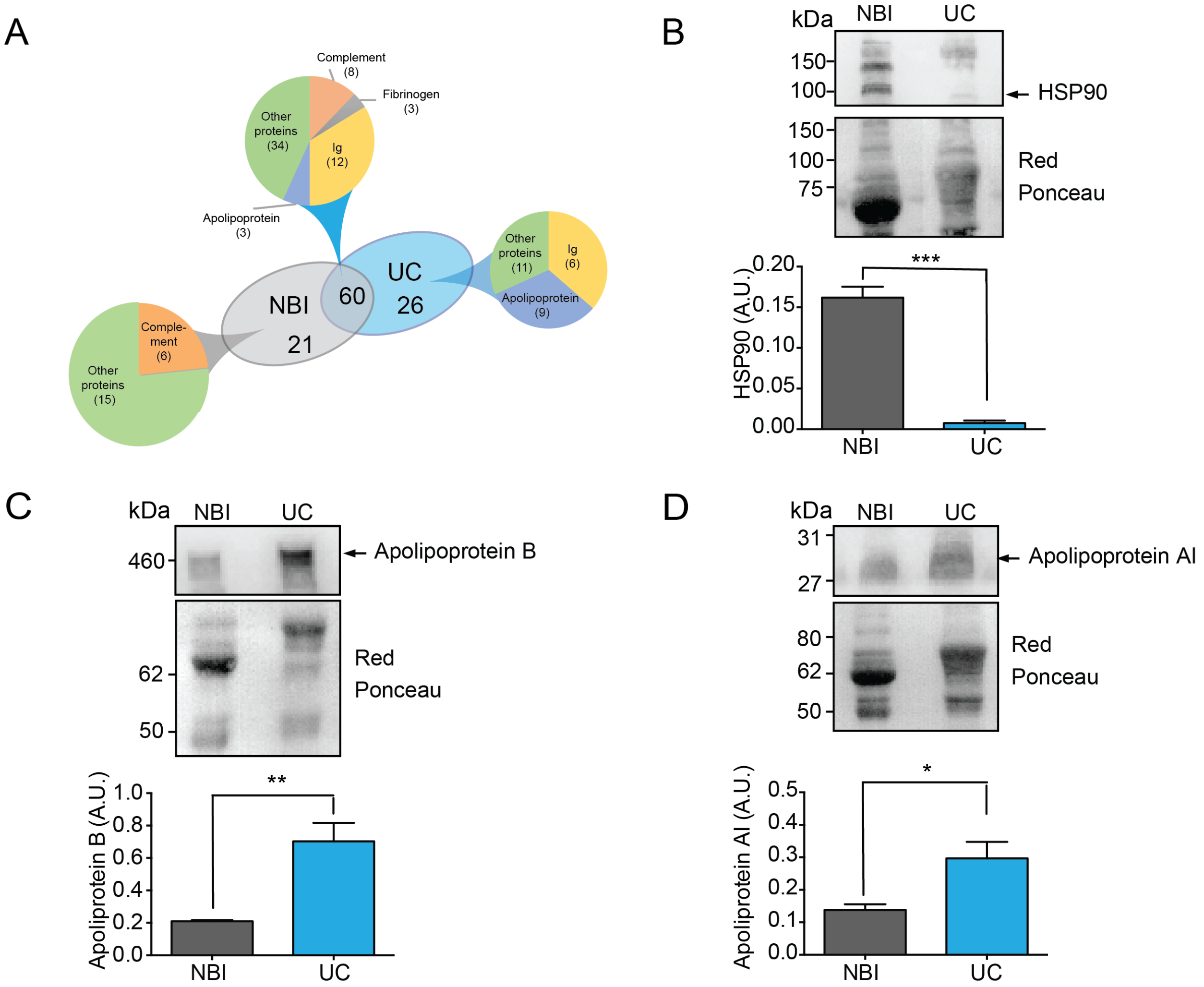


**Supplementary Figure 2.** EVs isolated with classical ultracentrifugation (UC) enriches for apolipoproteins than NBI. **a** Venn diagram of the unique and shared EV identified proteins isolated by NBI and UC methods. Numbers in brackets refer to the number of proteins belonging to each class. **b-d** Levels of HSP90 (**b**), Apolipoprotein B (**c**) and Apolipoprotein AI (**d**) in EVs isolated by NBI and UC methods (n=3). Student t-test; ***p=0.0002 for HSP90; **p=0.0063 for Apolipoprotein B; *p=0.0208 for Apolipoprotein AI. A.U.: arbitrary units.

Supplementary Figure 3

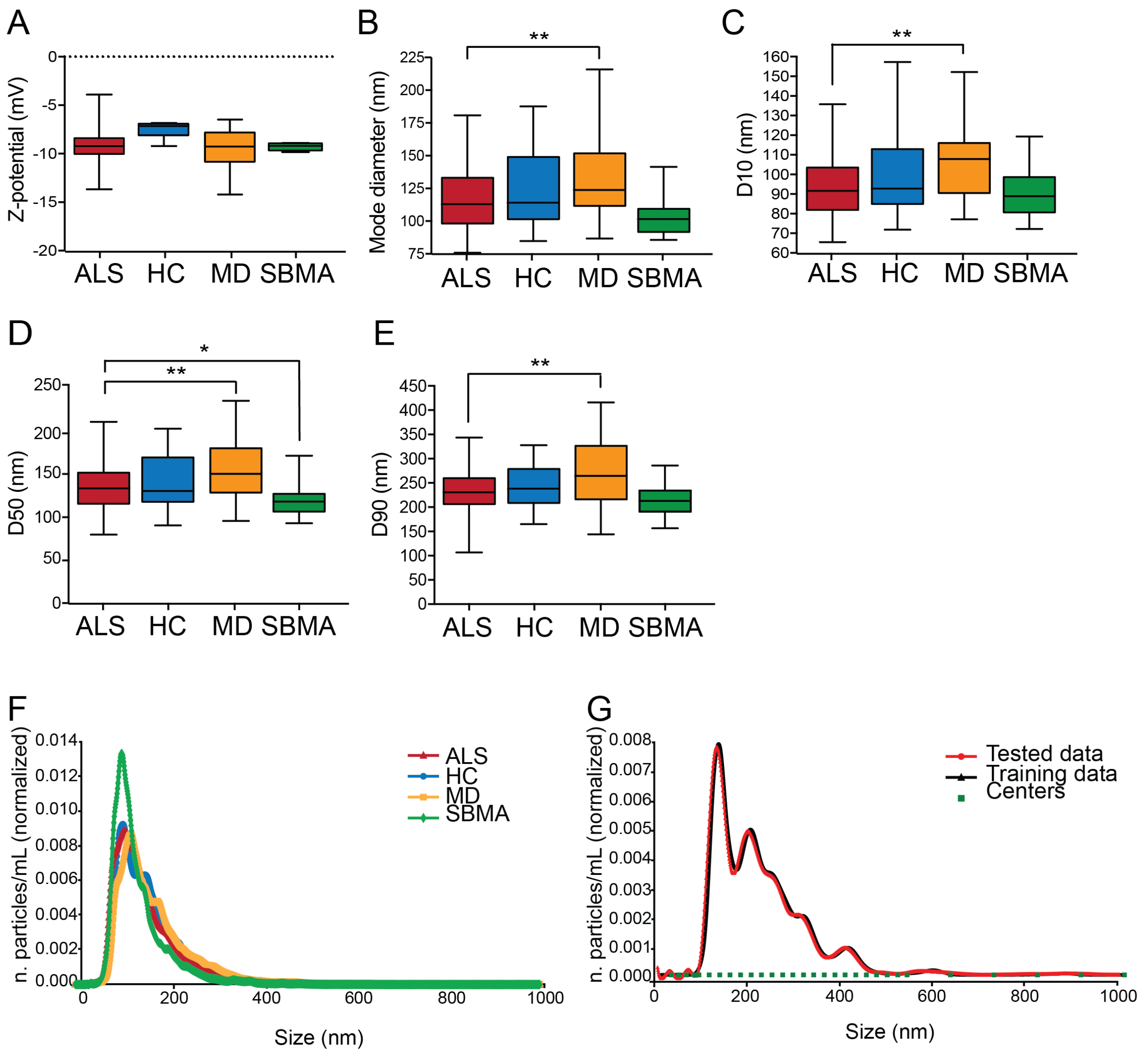


**Supplementary Figure 3.** Zeta potential and additional size parameters for EV plasma of ALS, HC, MD and SBMA. **a** Box plot for the average zeta potential (z-potential) for ALS, HC, MD and SBMA. **b** Box plot presenting the mode diameter (nm) of EVs purified from ALS, HC, MD and SBMA plasma. One-way ANOVA, p=0.0001; **p=0.0042 between ALS and MD by Dunnett's multiple comparisons test. **c** Box plot showing the D10 (nm) of EVs purified from ALS, HC, MD and SBMA plasma. One-way ANOVA, p=0.0002; **p=0.001 between ALS and MD by Dunnett's multiple comparisons test. **d** Box plot showing the D50 (nm) of EVs purified from ALS, HC, MD and SBMA plasma. One-way ANOVA, p<0.0001; **p=0.002 between ALS and MD, *p=0.021 between ALS and SBMA by Dunnett's multiple comparisons test. **e** Box plot showing the D90 (nm) of EVs purified from ALS, HC, MD and SBMA plasma. One-way ANOVA, p<0.0001; **p=0.0014 between ALS and MD by Dunnett's multiple comparisons test. **f** Normalized curves. **g** Representative image of the compressed size distribution using RBF. The green points are selected center, the black line is the initial signal, and the red line is the RBF-output signal.

Supplementary Figure 4

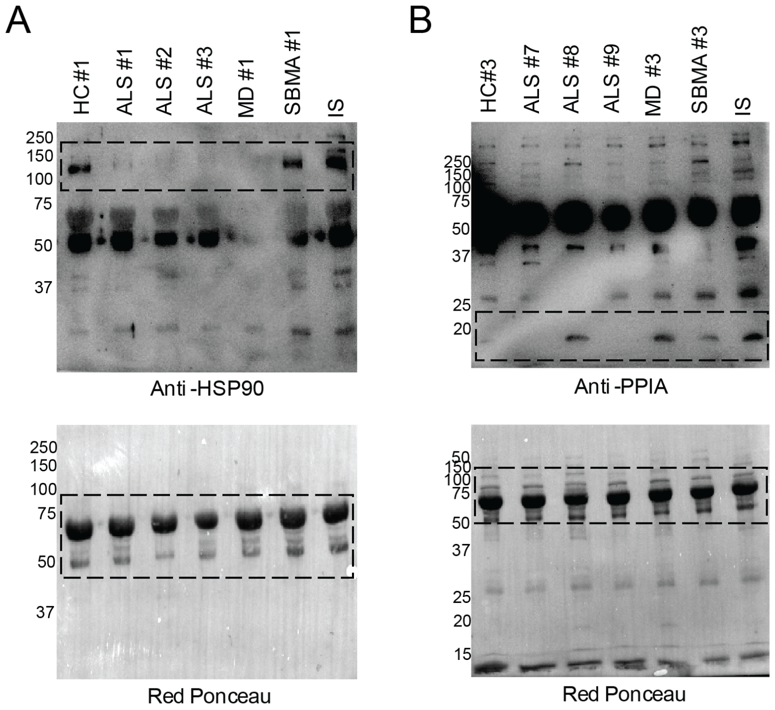


**Supplementary Figure 4. a, b** Complete immunoblotting for HSP90 (**a**), and PPIA (**b**) and relative Red Ponceau in human EV samples. IS means internal standard. The numbers appearing next to the different samples represent the actual number we assigned to each sample. The dashed box represents the blots reported in **Fig. 4**.

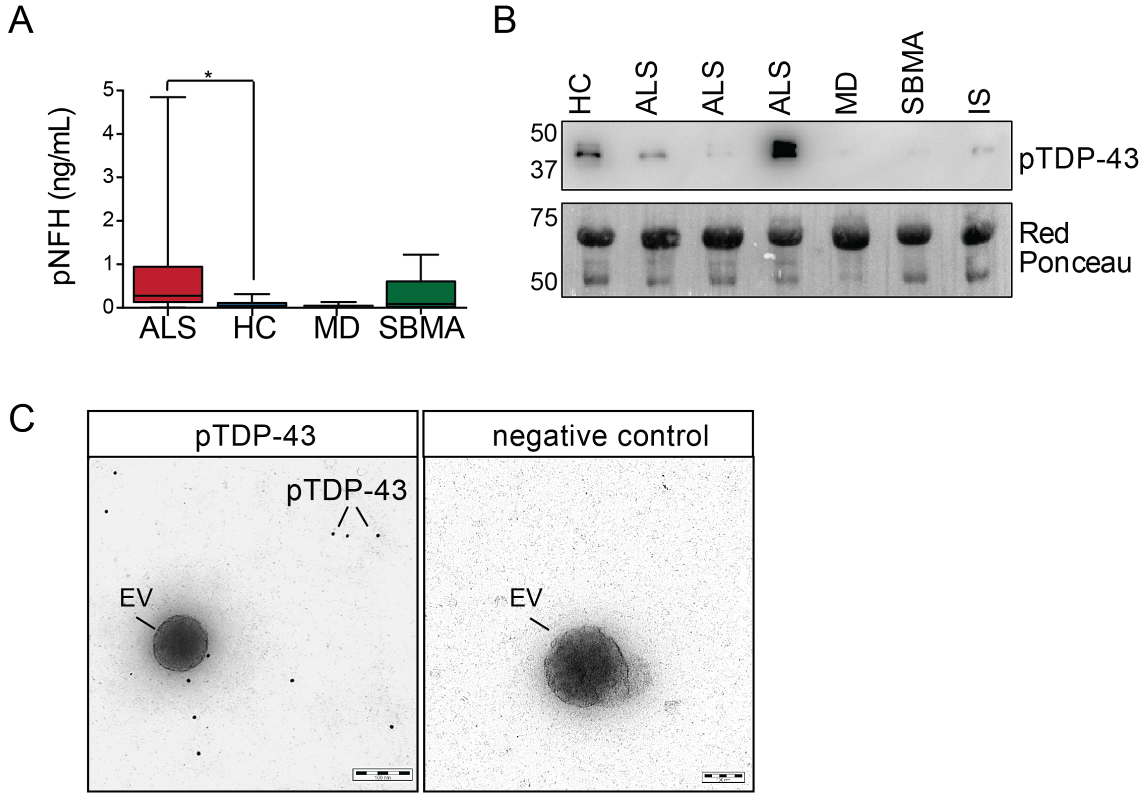


**Supplementary Figure 5.** Set up for the immunoblotting against TDP-43 and phosphorylated TDP-43, representative experiments. **a** Immunoblotting for anti-TDP-43 (C-terminus) in ALS. **b** Immunoblotting for anti-TDP-43 (N-terminus) in ALS samples. **c** Immunoblotting for anti-phosphorylated TDP-43 (pTDP-43) in ALS samples. **d** Red Ponceau relative to **a**, **b** and **c**. **e** Immunoblotting for anti- phosphorylated TDP-43 (pTDP-43) in human samples and relative Ponceau, referred to **Fig. 5b** (cropped area). **f** Immunogold transmission electron microscopy (TEM) analysis of plasma EVs purified with UC and stained with phosphorylated TDP-43 antibody (left panel) or negative control (right panel). Phosphorylated TDP-43 is indicated with a line (12 nm gold nanoparticles).
